# Usefulness of apparent diffusion coefficient values and magnetic resonance imaging histogram analysis for identifying histological types of preoperative testicular tumors

**DOI:** 10.1101/2024.11.12.24317155

**Authors:** Yuka Yasuda, Akiyoshi Osaka, Keita Izumi, Toshiyuki Iwahata, Akinori Nakayama, Kazunori Kubota, Kazutaka Saito

## Abstract

This study aimed to evaluate the ability of magnetic resonance imaging (MRI), including diffusion-weighted imaging with apparent diffusion coefficient values, to differentiate histological types of testicular tumors. Of 156 testicular tumors diagnosed at our hospital between January 2010 and July 2023, 65 cases with MRI were included. Tumors were categorized as seminoma, non-seminoma, and malignant lymphoma. Apparent diffusion coefficient values were calculated and analyzed using the ratio to normal testes and histograms according to tumor subtypes. Among the 65 cases, 46 were seminomas, 14 non-seminomas, and 5 malignant lymphomas. The apparent diffusion coefficient value ratio of seminomas was significantly higher than that of malignant lymphomas (p = 0.013). Additionally, the apparent diffusion coefficient value ratio of non-seminomas was significantly higher than that of seminomas and malignant lymphomas (p = 0.0013 and p < 0.001, respectively). On apparent diffusion coefficient histograms, malignant lymphomas had significantly higher values of kurtosis and skewness than seminomas. Normal testes and non-seminomas had significantly higher kurtosis and skewness values than that of malignant lymphomas and seminomas. A scoring model for seminomas and malignant lymphomas, developed with age and apparent diffusion coefficient value ratios, discriminated against the probability of seminoma and malignant lymphomas. MRI using apparent diffusion coefficient values and histogram analysis may aid the histological typing of seminomas, non-seminomas, malignant lymphomas.

## Introduction

Testicular tumors are heterogeneous and include various histologic subtypes, including seminoma and non-seminoma. Moreover, malignant lymphoma occurs in the testis without lymph node involvement [1]. While histological subtypes are determined through pathological evaluation, preoperative evaluation is crucial for developing appropriate treatment plans [2]. Imaging-based testicular tumor diagnosis for tumor presence and qualitative evaluation, including the tumor histological subtype, are essential. Magnetic resonance imaging (MRI) enables detailed internal anatomical and qualitative assessments and is useful in diagnostic imaging, which is widely used clinically [3–7]. However, detailed studies using MRI for the differential diagnosis of testicular tumor histological subtypes are lacking. Diffusion-weighted imaging (DWI) in MRI produces an apparent diffusion coefficient (ADC) map via ADC values and is useful in cancer diagnosis [8–11]. Tumor tissues demonstrate low ADC values owing to the restriction of free extracellular water molecule diffusion caused by increased cancer cell density and irregular growth [8]. Histogram analysis has attracted considerable attention in recent years. However, few studies have performed histogram analysis for the differential diagnosis of testicular tumors. We herein investigated the usefulness of MRI for testicular tumors using ADC values and histogram analysis for each testicular tumor histological subtype.

## Materials and methods

### Patients

This retrospective research has been approved by the Bioethics Committee of Dokkyo Medical University Saitama Medical Center (approval number: 24007). The university’s website has published an information release regarding this study. No patients involved in the study cases requested to refuse participation. The Bioethics Committee waived the requirement for informed consent due to the retrospective nature of the study. The planned research period spans from March 21, 2024, to March 31, 2025. Data was accessed from March 21, 2024, to October 26, 2024. During data collection, the authors had access to information that could identify individual participants. Of the 156 consecutive testicular tumors diagnosed at our hospital between January 2010 and July 2023, MRI with DWI performed before testicular resection was included in 77 cases. Among them, 69 cases were diagnosed as malignant testicular tumors, of which four cases of the single testis after orchiectomy were excluded to compare tumor testes and contralateral normal testes in the same patients. Cases with MRI but without DWI were excluded from the study. The remaining 65 cases were included in this study and classified as follows: seminomas, n = 46 cases; non-seminomas, n = 14; and malignant lymphomas, n = 5.

### MRI procedures and measuring ADC values

The MAGNETOM Avanto Dot Upgr 1.5T (SIEMENS, Munich, Germany), MAGNETOM Skyra 3.0T (SIEMENS), and Ingenia CX 3.0T (PHILIPS, Amsterdam, the Netherlands) MRI systems were used in this study. DWI was performed using echo-planar imaging with a b-value of 0 s/mm^2^ for Ingenia CX and 1,000 or 1,500 s/mm^2^ for MAGNETOM Avanto Dot Upgr and MAGNETOM Skyra. As three different ADC measurement procedures were used in this study, the ADC values of normal testes differed between them. To account for this difference, the ADC value ratio of tumor testis to normal testis across all three procedures was calculated. Normal testes were defined as testes with no abnormal findings on MRI imaging. The mean ADC values of tumor and normal testes were measured by setting a region of interest (ROI) on the largest cross-section of the testis on the ADC map, regardless of the presence of hemorrhage or necrotic lesions as it is difficult to distinguish them from tumor tissue on MRI images. Histogram analysis was performed to measure kurtosis and skewness.

### Statistical analysis

Data are presented as mean ± standard deviation. Significant differences were determined using the Wilcoxon test, and multiple comparisons were conducted using the Scheffe method. To distinguish testicular malignant lymphomas from seminomas, the score model was performed with the age at diagnosis and ADC values. It has been reported that the typical onset ages of malignant lymphoma are higher—typically in the 50s and 60s, compared to those of seminomas, which commonly occur in the 30s to 40s [12,13]. The total scores for seminomas and malignant lymphomas were calculated by adding their ADC value ratios. An ROC analysis was performed with a cut-off value of 0.637 for the ADC, with a specificity of 0.800 and a sensitivity of 0.804.

One point was given for ages < 30 years and > 50 years, 0 points for those aged 30–50 years of age, one point for the ADC value ratio of > 0.637, and 0 points for ≥ 0.637.

## Results

### Patient background characteristics

Patients’ age ranges were 24–59 (median, 38.7) years for seminomas, 19–51 (median, 33.6) years for non-seminomas, and 24–74 (median, 56.8) years for malignant lymphomas. The histological types for non-seminomas for each case are shown in Table 1. Of 14 non-seminoma patients, three patients had pure histology of embryonal carcinoma, germ cell tumor, or teratoma. The histology of the 11 remaining patients was mixed non-seminomas. All five testicular malignant lymphoma patients had diffuse large B-cell lymphoma histology.

**Table 1.** Patients’ background characteristics.

|  | Seminoma | Non-seminoma | Malignant lymphoma |
| --- | --- | --- | --- |
| Number of cases | 46 | 14 | 5 |
| Age (years) | 24–59 (38.7) | 19–51 (33.6) | 24–74 (56.8) |
| T1WI | ↓ | ↓ | ↓ |
| T2WI | ↓ | ↑+↓ | ↓ |
| DWI | ↑ | ↑+↓ | ↑ |
| ADC | ↓ | ↑+↓ | ↓ |
| ADC values ratio | 0.745±0.132 | 1.197±0.430 | 0.531±0.119 |
| Tumor size (mm<sup>2</sup>) | 1133±973 | 1705±1605 | 1557±962 |

Data is presented as range (mean).
High signal: ↑, low signal: ↓

### Testicular MRI findings

Normal testes showed low signal intensity on T1-weighted imaging (T1WI) and had high ADC values on MRI. In contrast, seminomas and malignant lymphomas showed high signal intensity on T2-weighted imaging (T2WI) and DWI (Fig 1 and 2). Non-seminomas often contained multiple tissues and did not show a single signal intensity (Fig 3). However, seminomas and malignant lymphomas had similar signal intensities, making it difficult to distinguish them using T2WI or DWI (Fig 1 and 2).

**Fig 1.** Representative findings of seminoma. T1W1 (a), T2W1 (b), DWI (c), and ADC map (d). The ROI surrounded the entire tumor on the ADC map (e). Histogram of ADC value (f). The histogram of the seminoma showed high kurtosis and skewness values of 15.318 and 3.21, respectively. It showed the distribution with peaked and heavy fringes compared with the normal distribution, and the mean ADC values were skewed toward the lower end of the distribution.

**Fig 2.** Representative findings of malignant lymphoma. T1W1(a), T2W1(b), DWI (c), and ADC map (d). The ROI surrounded the entire tumor on the ADC map (e). Histogram of the ADC value (f). It had a shape with peaked peaks and heavy fringes compared with the normal distribution. Furthermore, the overall distribution of ADC values was skewed toward lower values compared with that of seminoma.

**Fig 3.** Representative findings of non-seminoma. T1W1(a), T2W1(b), DWI (c), and ADC map (d). The ROI surrounded the entire tumor region on the ADC map (e). Histogram of the ADC value (f). The histogram of the non-seminoma showed low kurtosis and skewness values of 2.632 and 0.464, respectively. It showed a heavy hem, the distribution was symmetrically close, and the mean ADC values showed a broad distribution.

### ADC value ratio

The ADC value ratio was 0.745 ± 0.132, 1.197 ± 0.430, and 0.531 ± 0.119 for seminomas, non-seminomas, and malignant lymphomas, respectively (Table 1). The ADC value ratio of seminomas was significantly higher than those of malignant lymphomas (p = 0.013), and the ADC value ratios of non-seminomas were significantly higher than those of seminomas and malignant lymphomas (p = 0.0013 and p < 0.001, respectively) (Fig 4).

**Fig 4.** Distributions of the ADC value ratio.

### The ADC histogram: kurtosis and skewness

From the ADC histograms, kurtosis and skewness were measured. Kurtosis indicates the sharpness or heaviness of a data distribution base, and its value is high when the data are concentrated around the mean. Skewness indicates the degree of distribution asymmetry and is high when the ADC value is skewed toward the lower side [8]. The kurtosis and skewness of normal and each tumor testis were compared, although the shooting conditions are different. Seminomas and malignant lymphomas had significantly higher kurtosis values (8.55 ± 5.76 and 18.11 ± 5.22, respectively) compared with normal testes (6.20 ± 5.02, p = 0.0267 and p = 0.0044, respectively) and non-seminoma (4.92 ± 3.85, p = 0.012 and p = 0.0022, respectively). Malignant lymphomas had significantly higher kurtosis values than those of seminomas (p = 0.0123). No differences were observed between normal testes and non-seminomas (p = 0.3201). Seminomas and malignant lymphomas had significantly higher skewness values (1.77 ± 1.00 and 3.12 ± 0.28, respectively) compared with normal testes (0.50 ± 1.24, p < 0.001 and p < 0.001, respectively) and non-seminomas (0.52 ± 1.17, p = 0.0016 and p < 0.001, respectively). Malignant lymphomas had higher skewness than that of seminomas (p < 0.001). No differences were observed between normal testes and non-seminomas (p = 0.8907) (Fig 5a–c).

**Fig 5.** Skewness (a) and kurtosis (b) of the ADC value histogram. Correlation graph of skewness and kurtosis of the ADC value histogram (c).

### Scoring of seminomas and malignant lymphomas

Using the scoring model with ADC value ratio and age, 34 (73.9%), 10 (21.7%), and two (4.3%) patients with seminoma scored 0, 1, and 2 points, respectively (Figure 6a). In contrast, zero (0%), two (40%) and three (60%) patients with malignant lymphoma scored 0, 1, and 2, respectively (Figure 6b). In this scoring model, patients with scores of 0 and 1 have a high probability of seminoma, whereas those with a score of ≥ 2 have a high probability of malignant seminoma.

**Fig 6.** Probability of seminoma (a) and malignant lymphoma (b) according to the score.

## Discussion

This study indicated that MRI with ADC values and histograms are useful for characterizing testicular tumor subtypes. To our knowledge, this study is the first to demonstrate the significance of ADC values with histogram analysis in testicular tumor histological typing. MRI provides detailed internal anatomical and qualitative evaluations. Moreover, DWI presents biological tissue properties using the water molecules diffusion phenomenon and ADC values as parameters to evaluate the diffusion limit in vivo quantitatively [8–11]. It reflects tumor cell density, and its usefulness has been reported at various tumor sites [12,13]. We investigated the significance of ADC values in the histological typing of testicular tumor using the ADC value ratio of tumor testis to normal testis because three different procedures were used in ADC measurement during the study period. The current results demonstrate that seminomas and malignant lymphomas had lower ADC values than normal testes, and malignant lymphomas had even lower ADC value ratios than seminomas. Non-seminomas display a wide distribution of ADC value ratios because they often contain various histological types, and the ADC value ratios of each tissue type are different, as expected. Histogram analysis has attracted considerable attention in recent years. Moreover, ADC enables detailed tumor heterogeneity and cell density evaluation and aids histological diagnosis [14–19]. The statistical indices representing the distribution characteristics were kurtosis and skewness. In the ADC histogram analysis, kurtosis and skewness were high in seminomas and malignant lymphomas. The specific ADC values of seminomas and malignant lymphomas were prominent, and kurtosis was high, which could reflect cellular uniformity, unlike non-seminomas. Moreover, compared with normal testes, seminomas and malignant lymphomas showed low ADC values, which may have resulted in high skewness. Kurtosis and skewness were low in non-seminomas. Non-seminomas often contain various histological types, and their ADC values are widely and gently distributed, suggesting that they tend to have lower kurtosis and skewness than other malignant tumors. Testicular tumors include various subtypes categorized into seminomas or non-seminomas. Furthermore, malignant lymphomas are found as testicular tumors [20]. Although the diagnosis is made pathologically after the testis is resected, preoperative evaluation could be essential and beneficial in developing the treatment plan earlier [2]. Non-seminomas often contain multiple histological types, including embryonal carcinomas, yolk sac tumors, choriocarcinomas, and teratomas, and it can be challenging to distinguish non-seminomas from seminomas or malignant lymphomas with a mixture of high and low signal areas on testicular MRI images [21]. On the histograms, kurtosis and skewness of normal testes and non-seminomas were lower than those of seminomas and malignant lymphomas, as expected. Although it is outside the scope of this study, a detailed analysis of each histological type would be warranted in the future. Preoperative differential diagnosis between seminoma and testicular malignant lymphoma is often complex. The current results demonstrate the distinct characteristics of these based on DWI findings. Noting that the ADC value ratio of seminomas and malignant lymphomas are lower than those of non-seminomas, we attempted to score them using age and ADC value ratio to simply distinguish them. Among patients with seminoma, 73.9% had the highest score (0 points) for the possibility of seminoma. Among the malignant lymphoma cases, 60% had the highest score (2 points) for the possibility of malignant lymphoma. Since all types included a case with a score of 1, it is considered necessary to study a larger sample size, especially for malignant lymphoma, and to reexamine the ADC ratio value, including its setting value. We confirmed significant differences among histological types by measuring the ADC value ratio, analyzing ADC histograms, and scoring seminomas and malignant lymphomas based on age and ADC value ratio. These values can be used as an indicator for MRI diagnosis in cases like ours, where the histological type is challenging to determine. Furthermore, the detailed understanding of imaging findings may further elucidate testicular tumor biology.

This study had some limitations. The number of patients in this study was small—46 had seminomas, 14 had non-seminomas, and five had malignant lymphomas. The small number of cases may have caused bias or variation in the ADC value ratio. Testicular tumors have an incidence rate of 3–10 per 100,000 men [22]. It is not easy to collect enough cases to perform an adequate analysis. Furthermore, the ROI setting was performed only by the author in this study. Although the measurements were taken at the largest section of the tumor to avoid errors, it is thought that the accuracy of the results will increase if ROIs are set at multiple locations or measurements are taken by multiple persons. The accuracy of the chi-square test for scoring seminomas and malignant lymphomas would also be improved by increasing the number of cases.

## Conclusion

The ADC value ratio of seminomas was significantly higher than that of malignant lymphomas. Additionally, the ADC value ratio of non-seminomas was significantly higher than that of seminomas and malignant lymphomas. On apparent diffusion coefficient histograms, malignant lymphomas had significantly higher values of kurtosis and skewness than that of seminomas. Normal testes and non-seminomas had significantly higher kurtosis and skewness values than those of malignant lymphomas and seminomas. In conclusion, ADC value ratio and MRI-based ADC histogram analysis are useful for distinguishing between seminomas, non-seminomas, and malignant lymphomas.

## Data Availability

All relevant data are within the manuscript and its Supporting Information files.

## Acknowledgment

None.

---

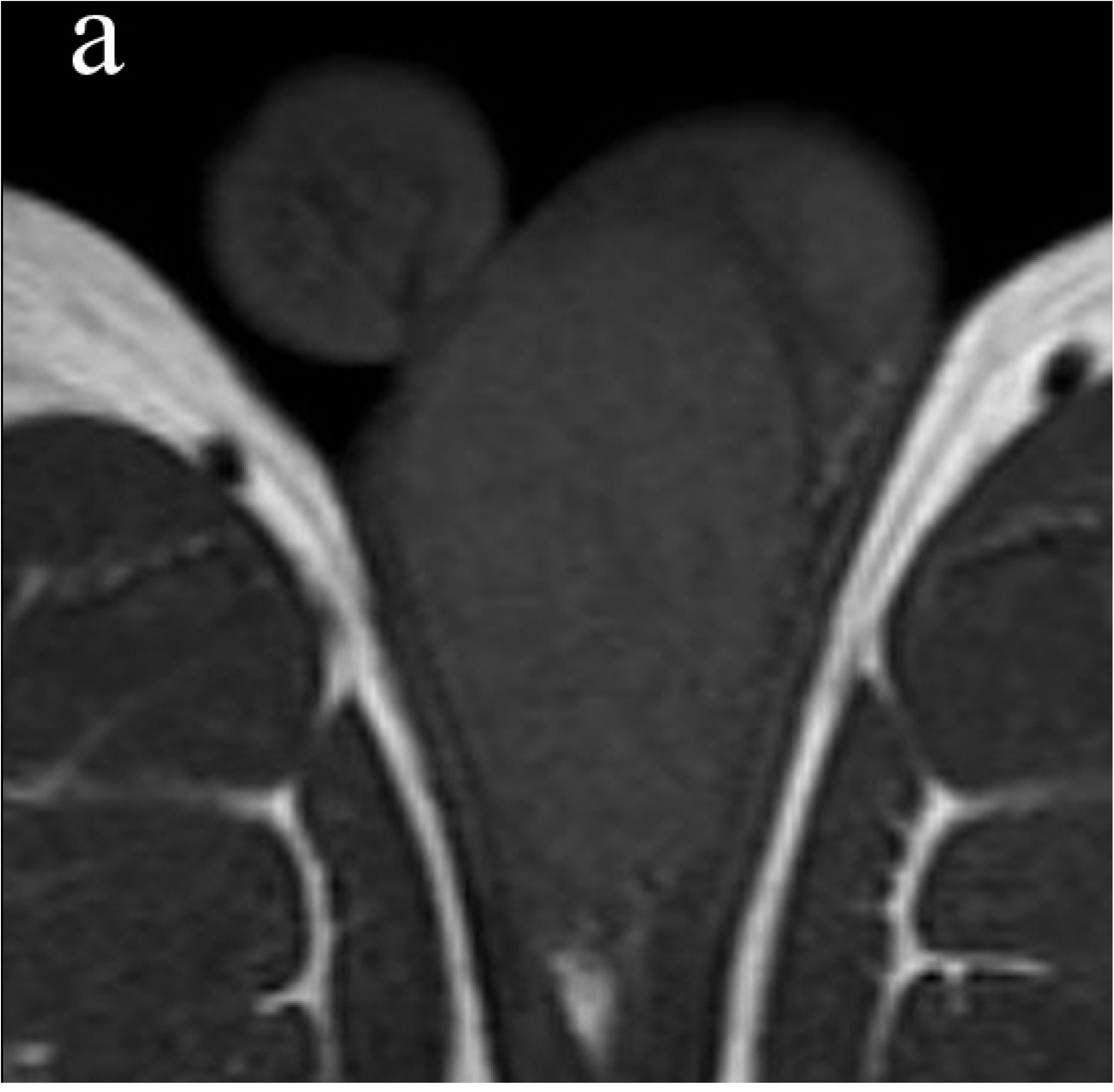

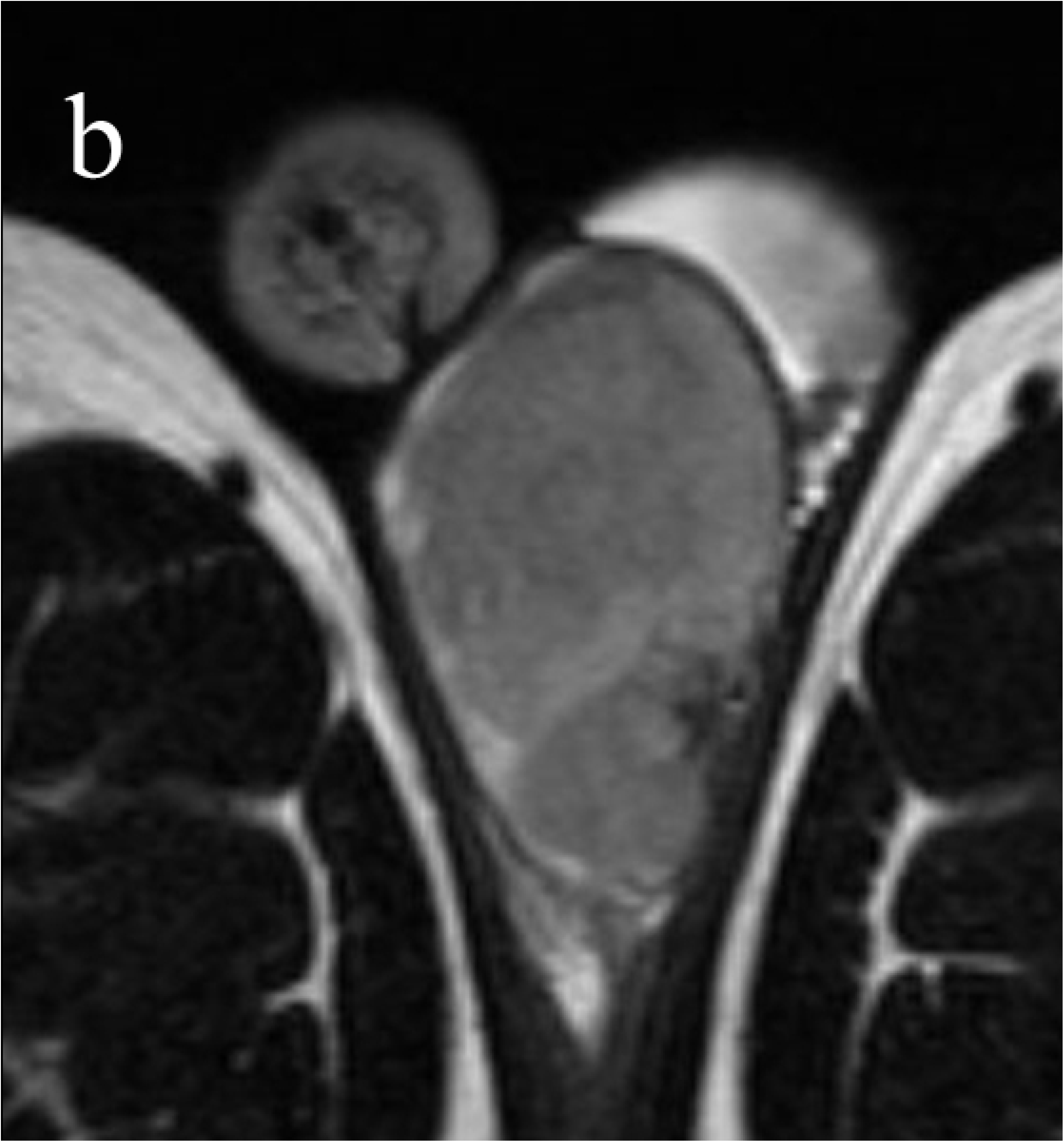

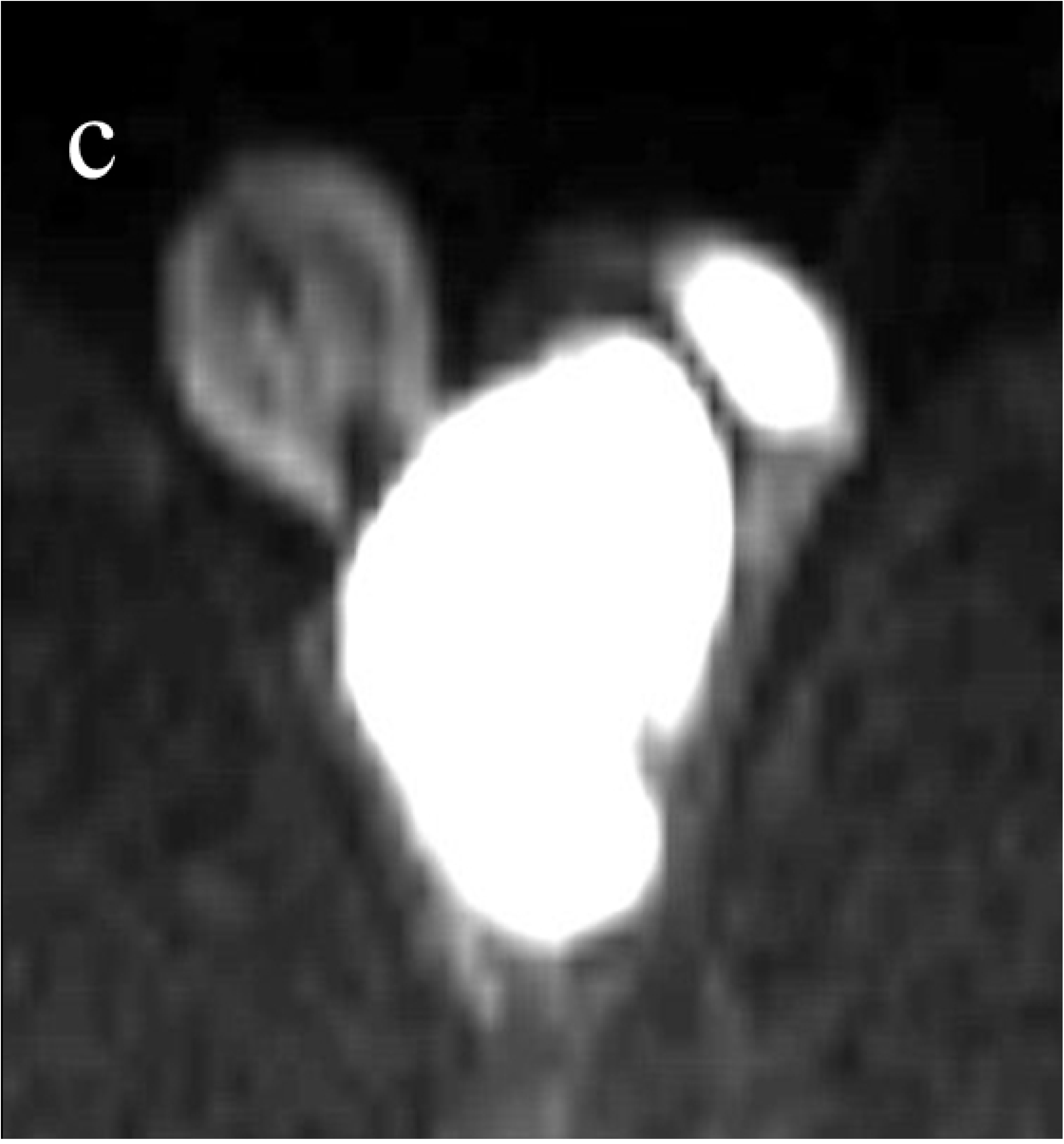

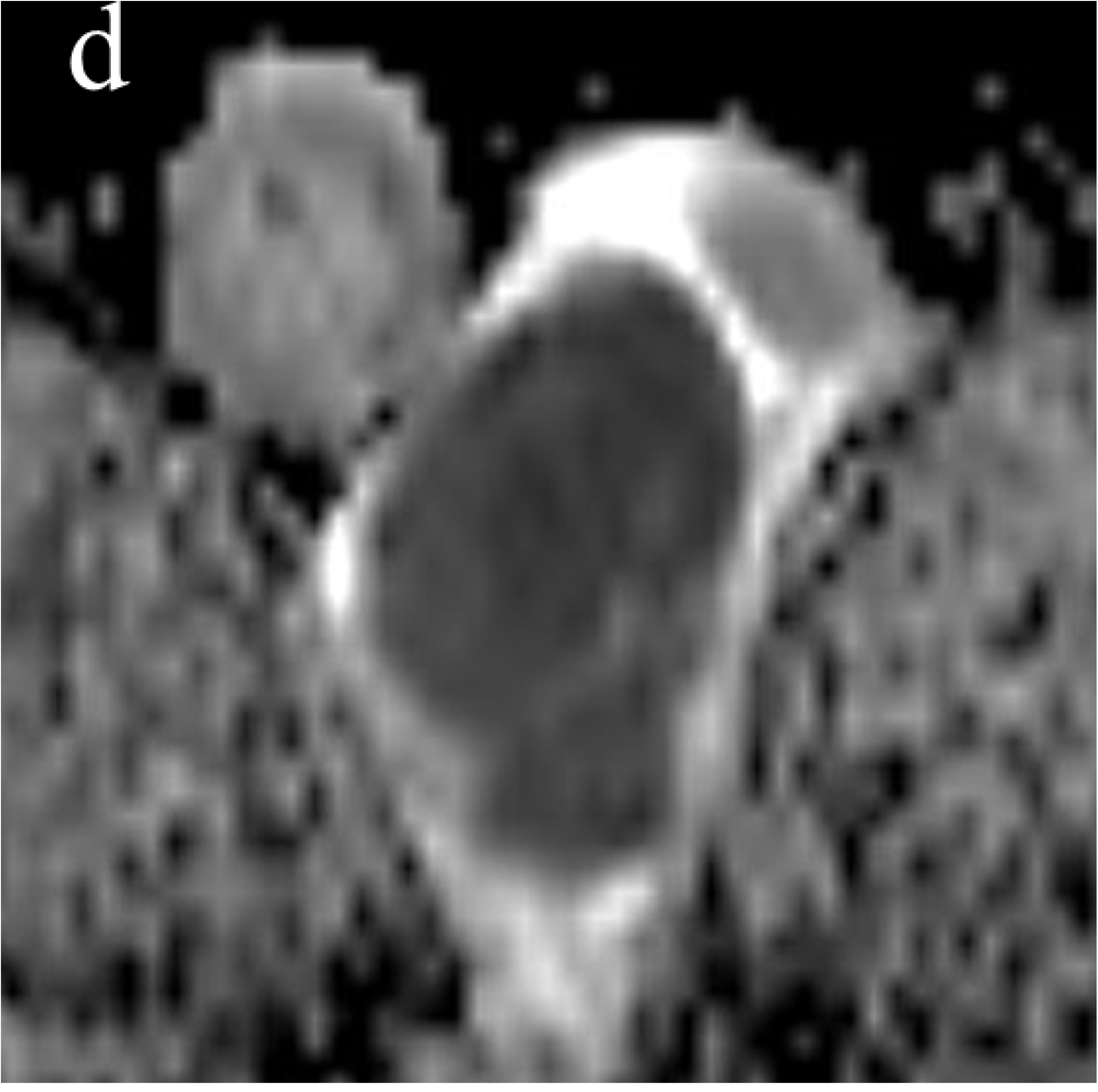

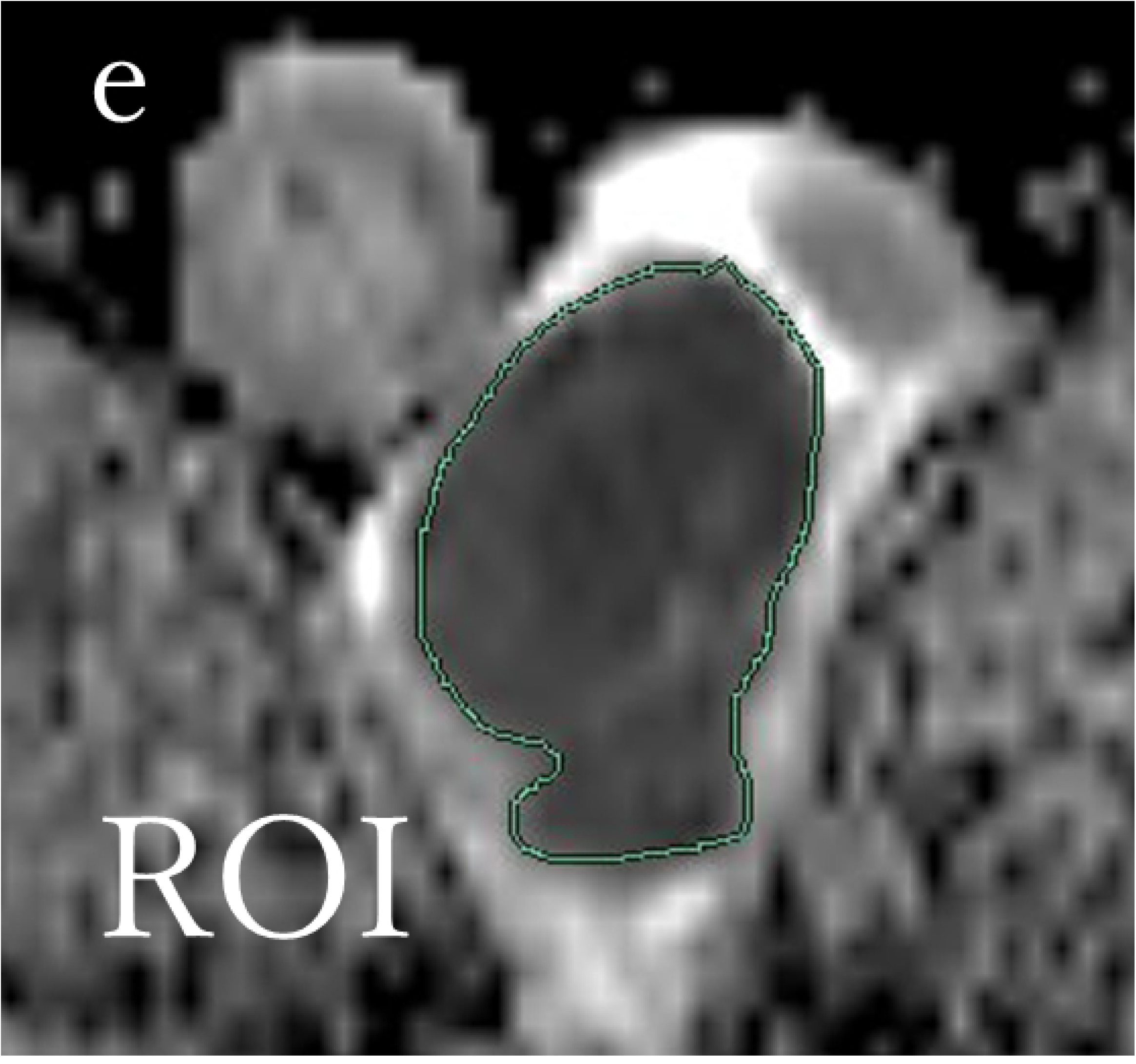

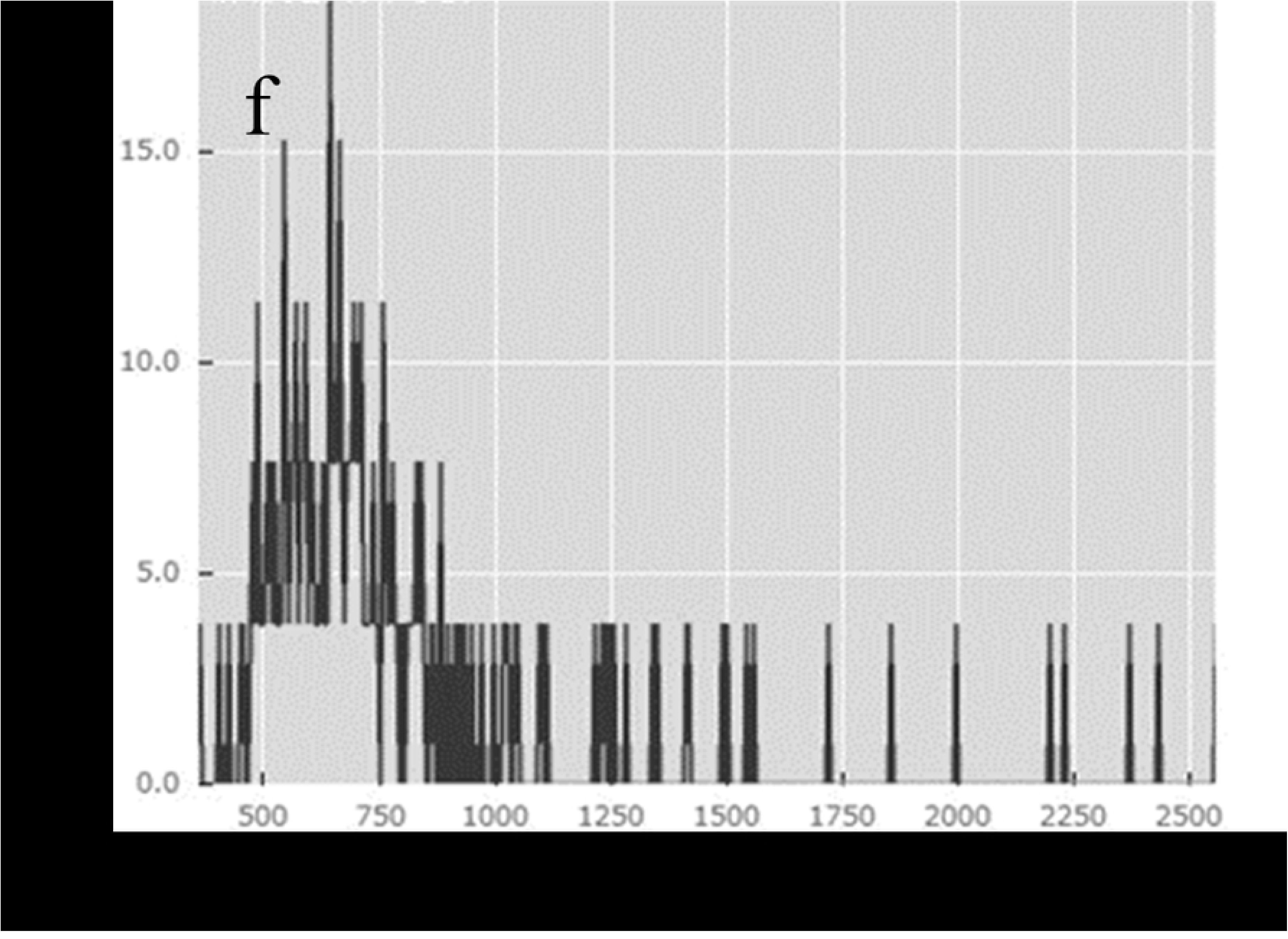

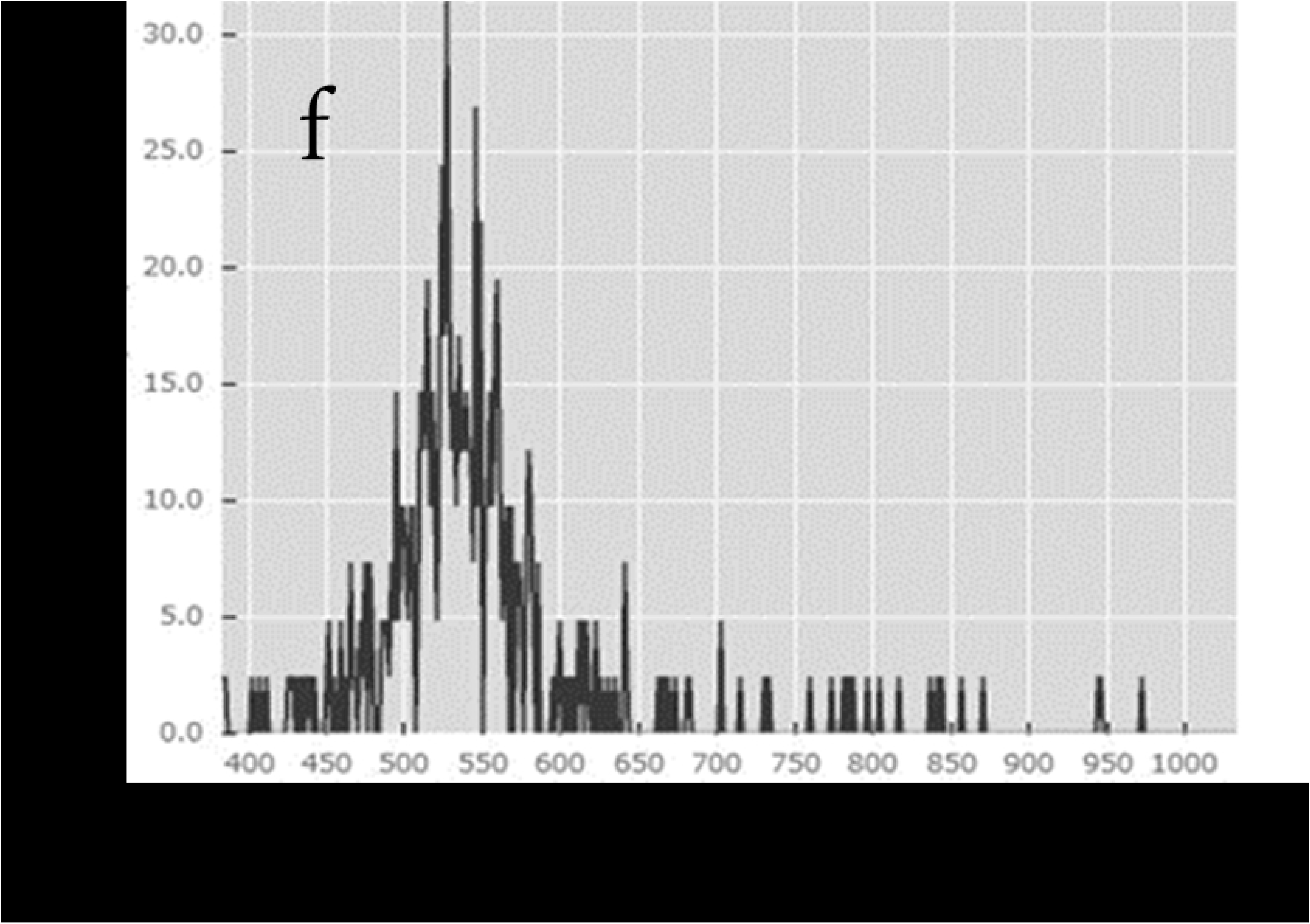

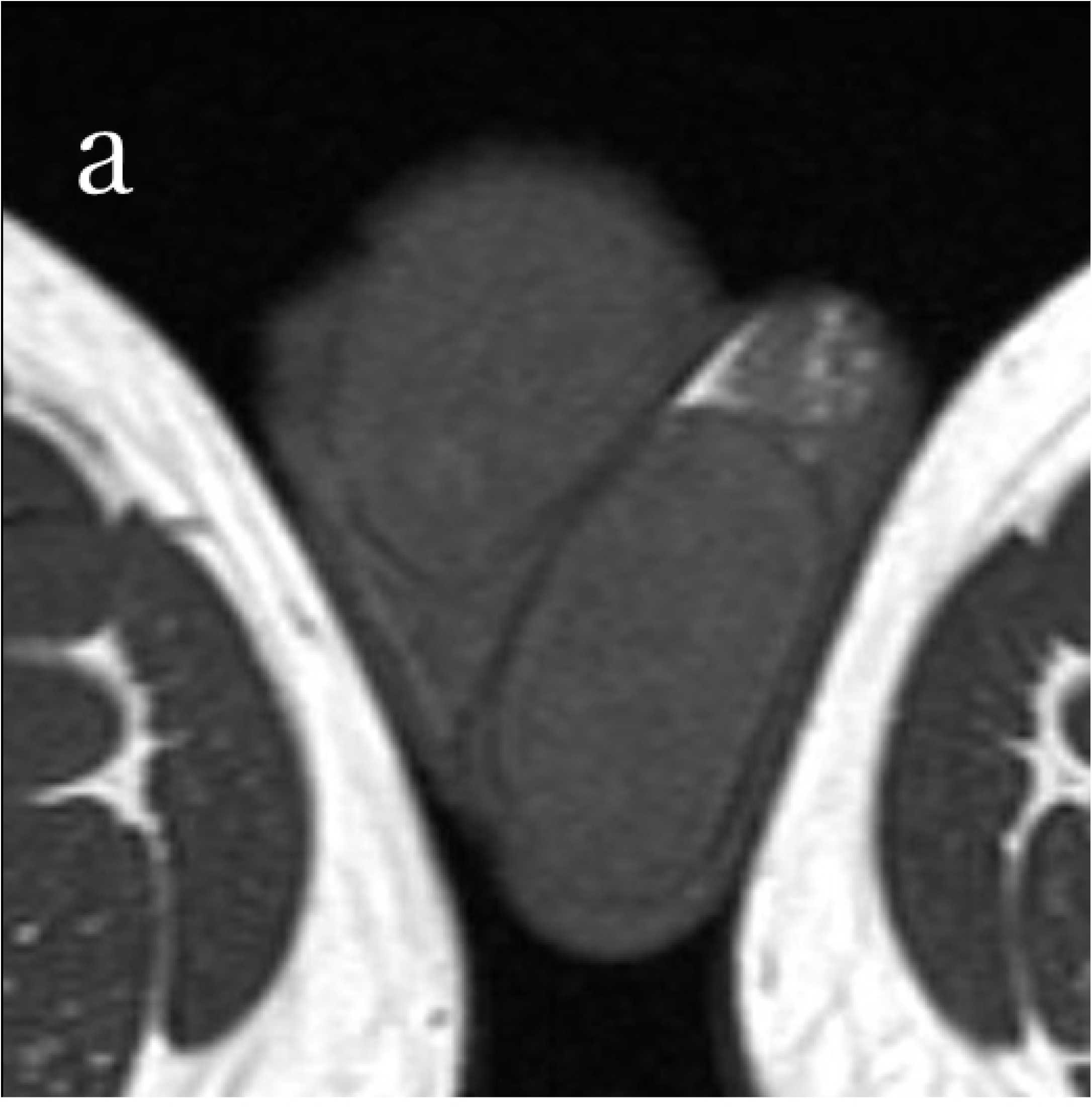

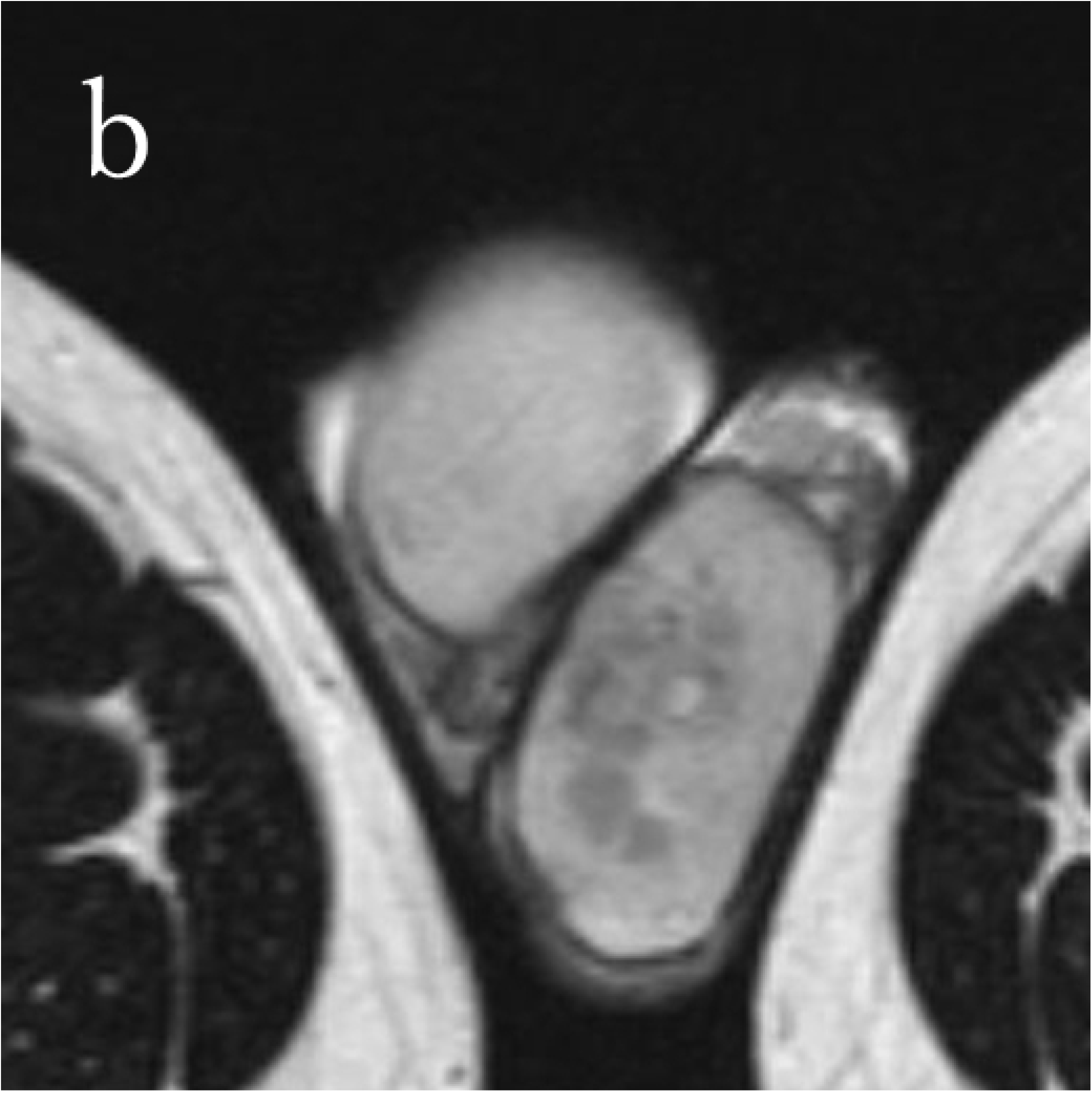

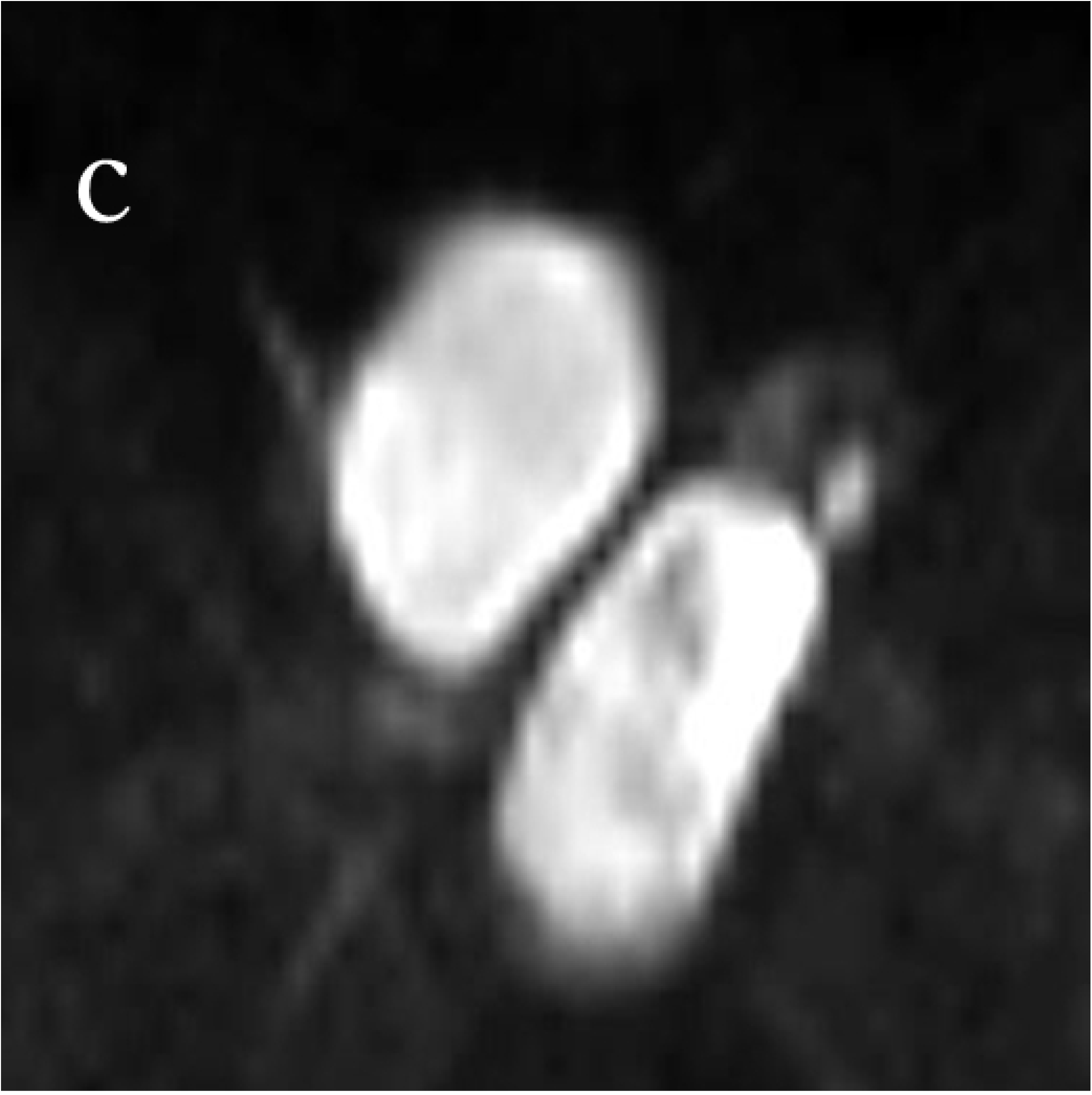

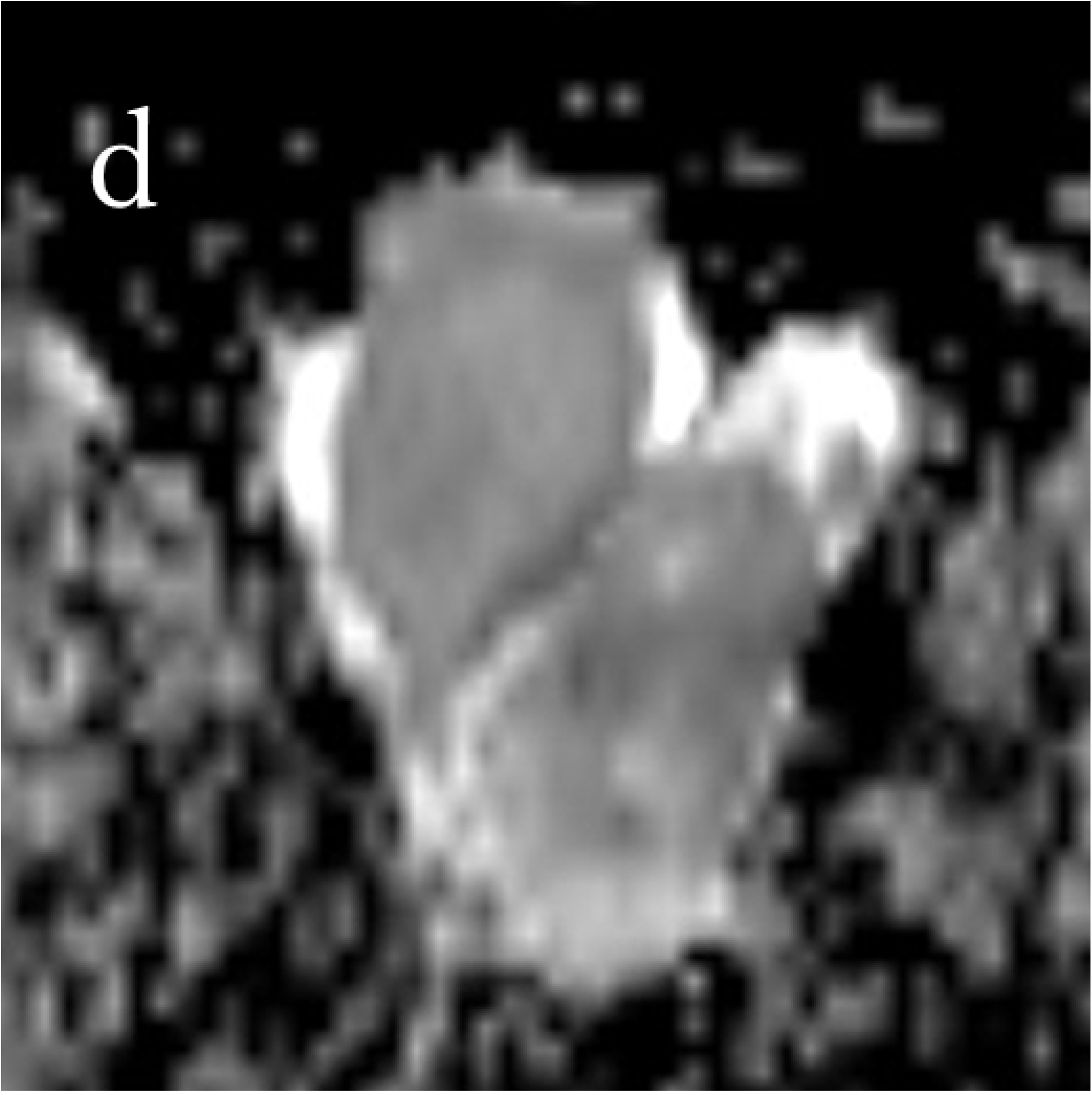

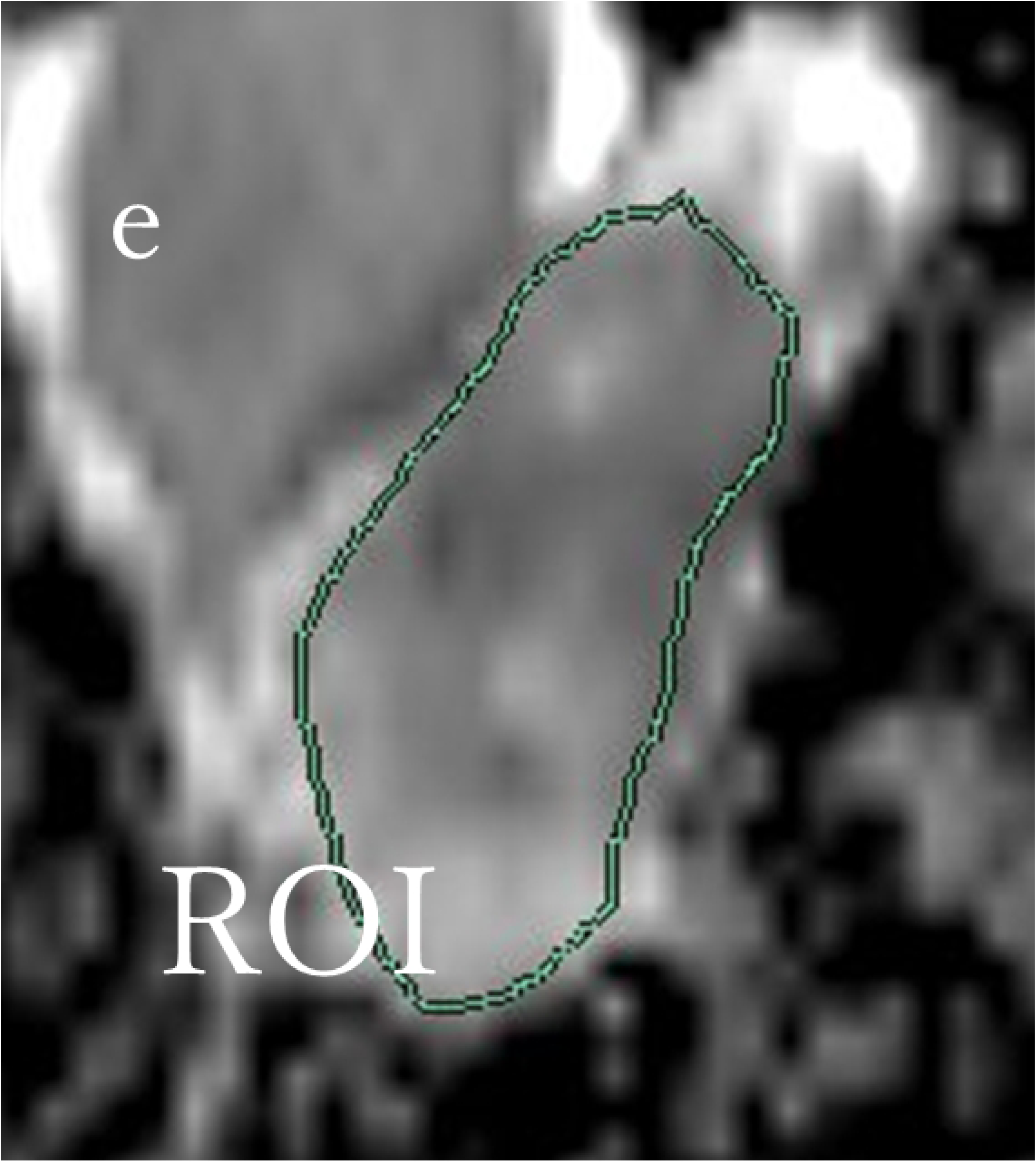

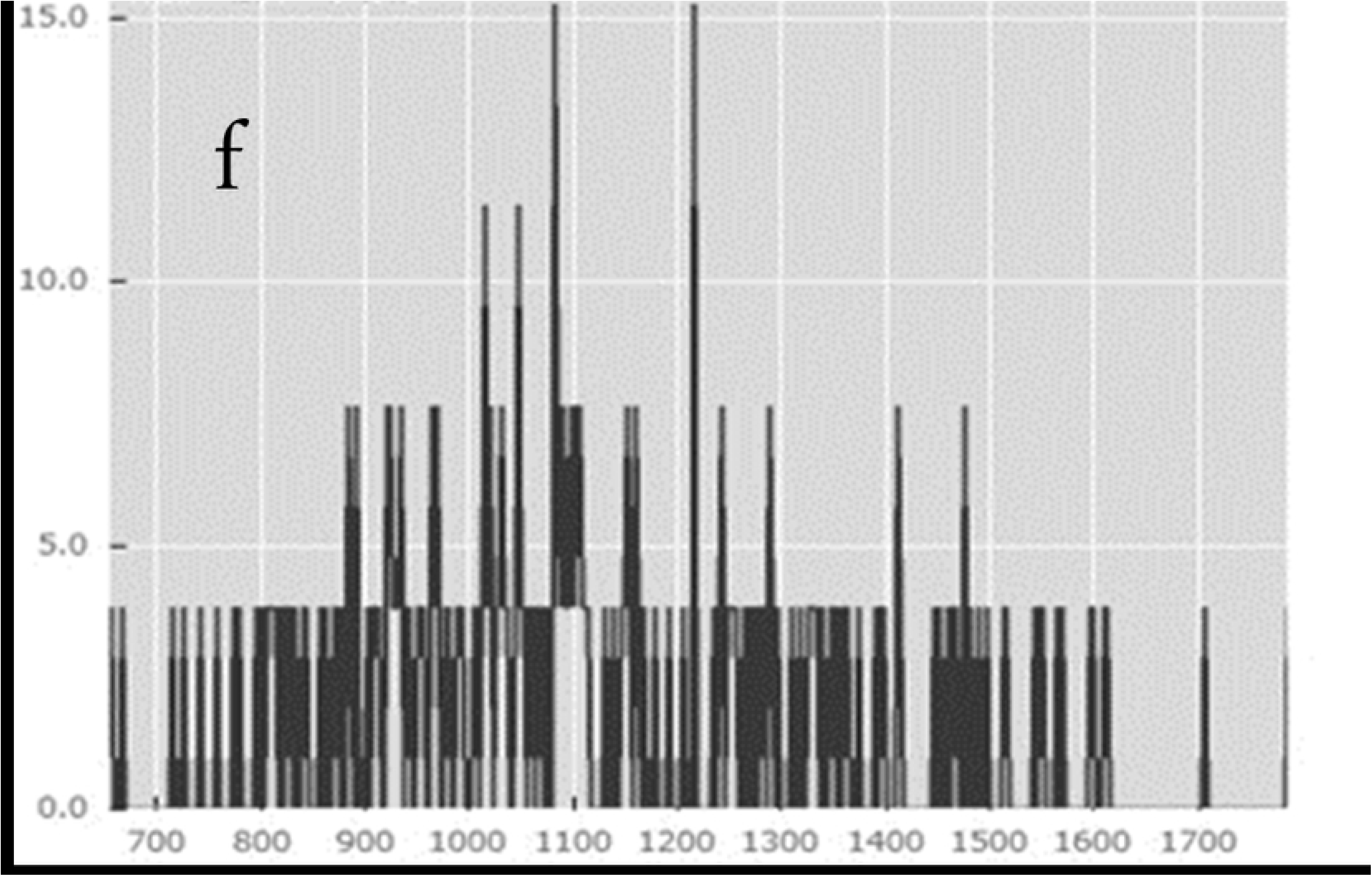

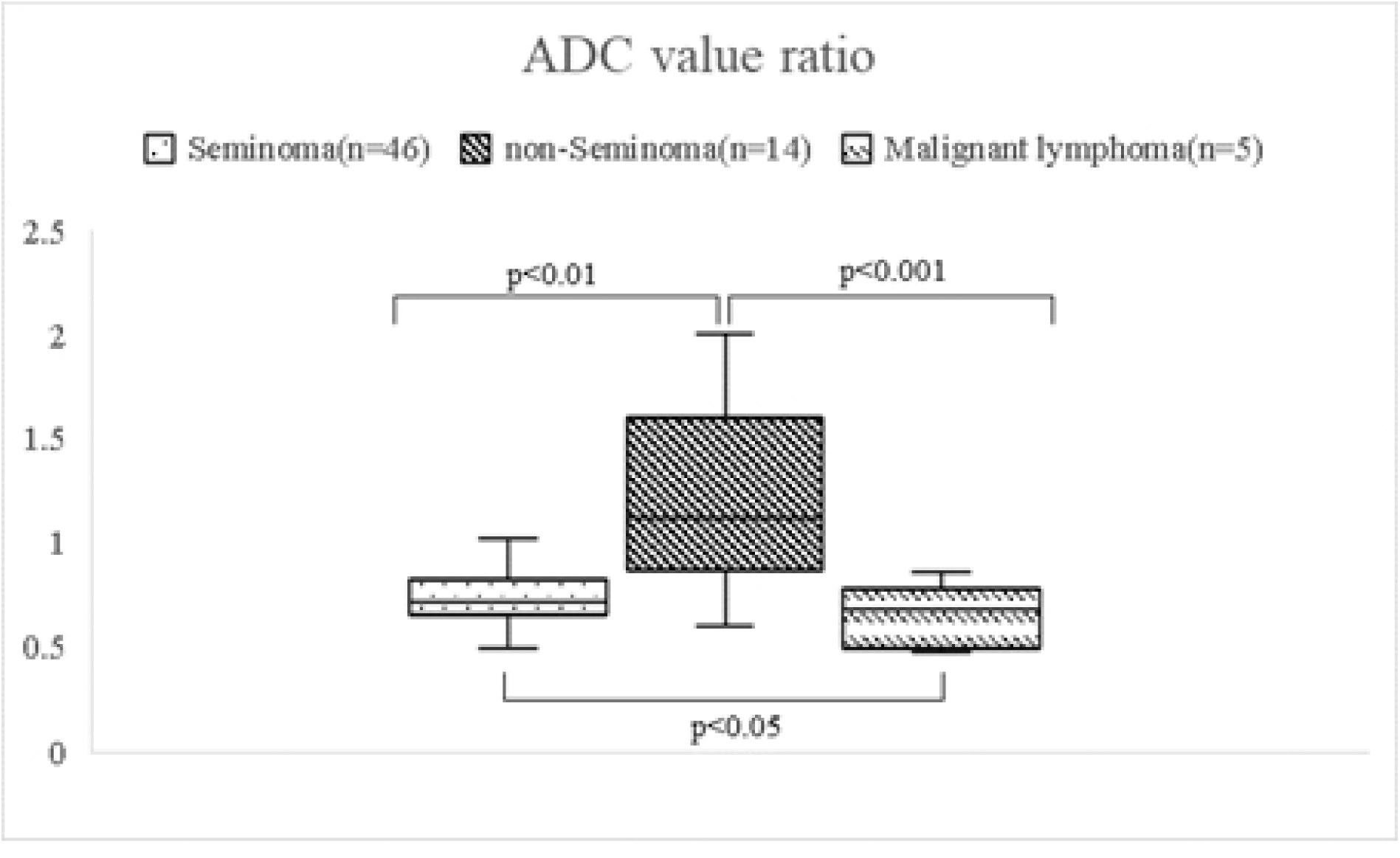

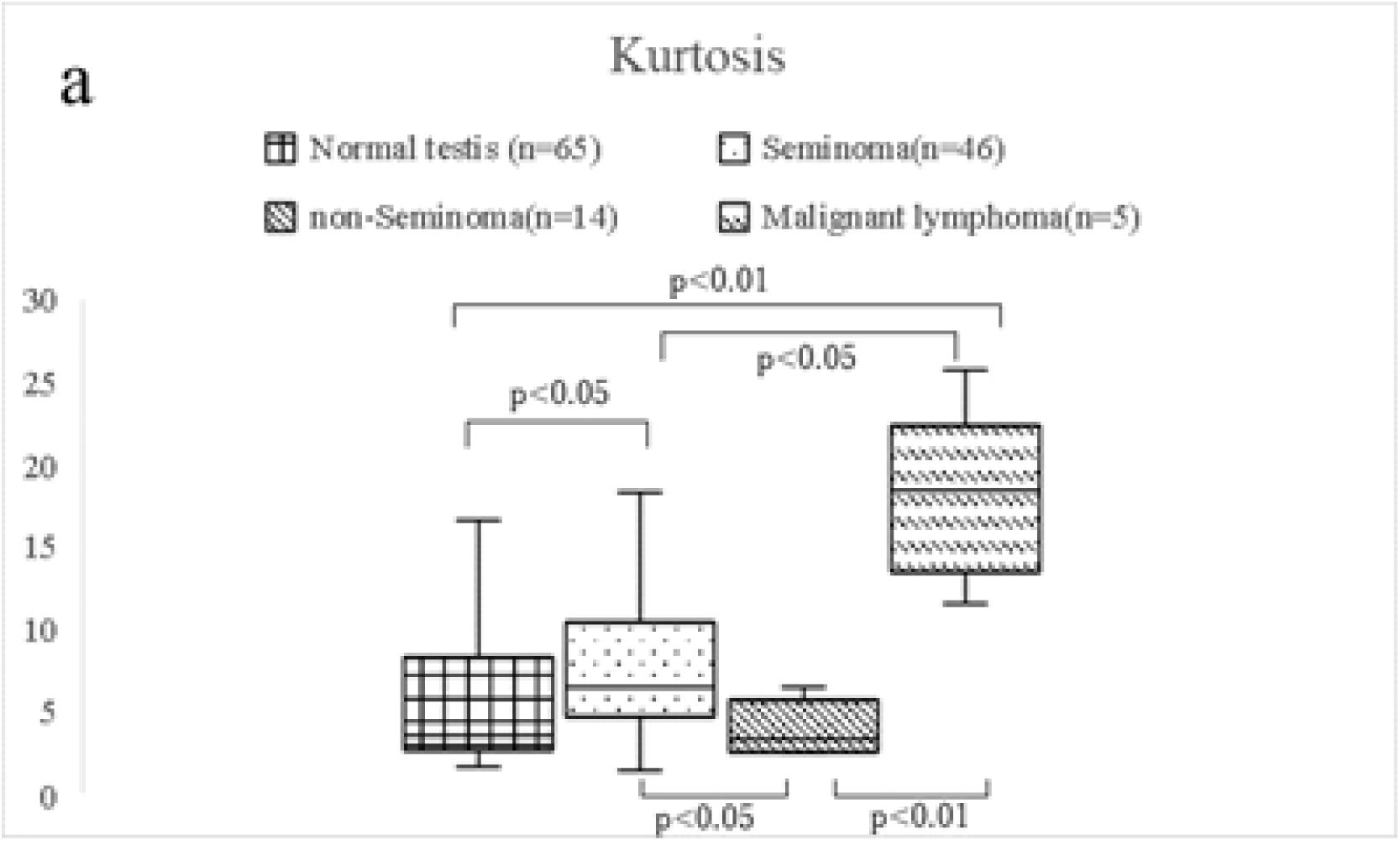

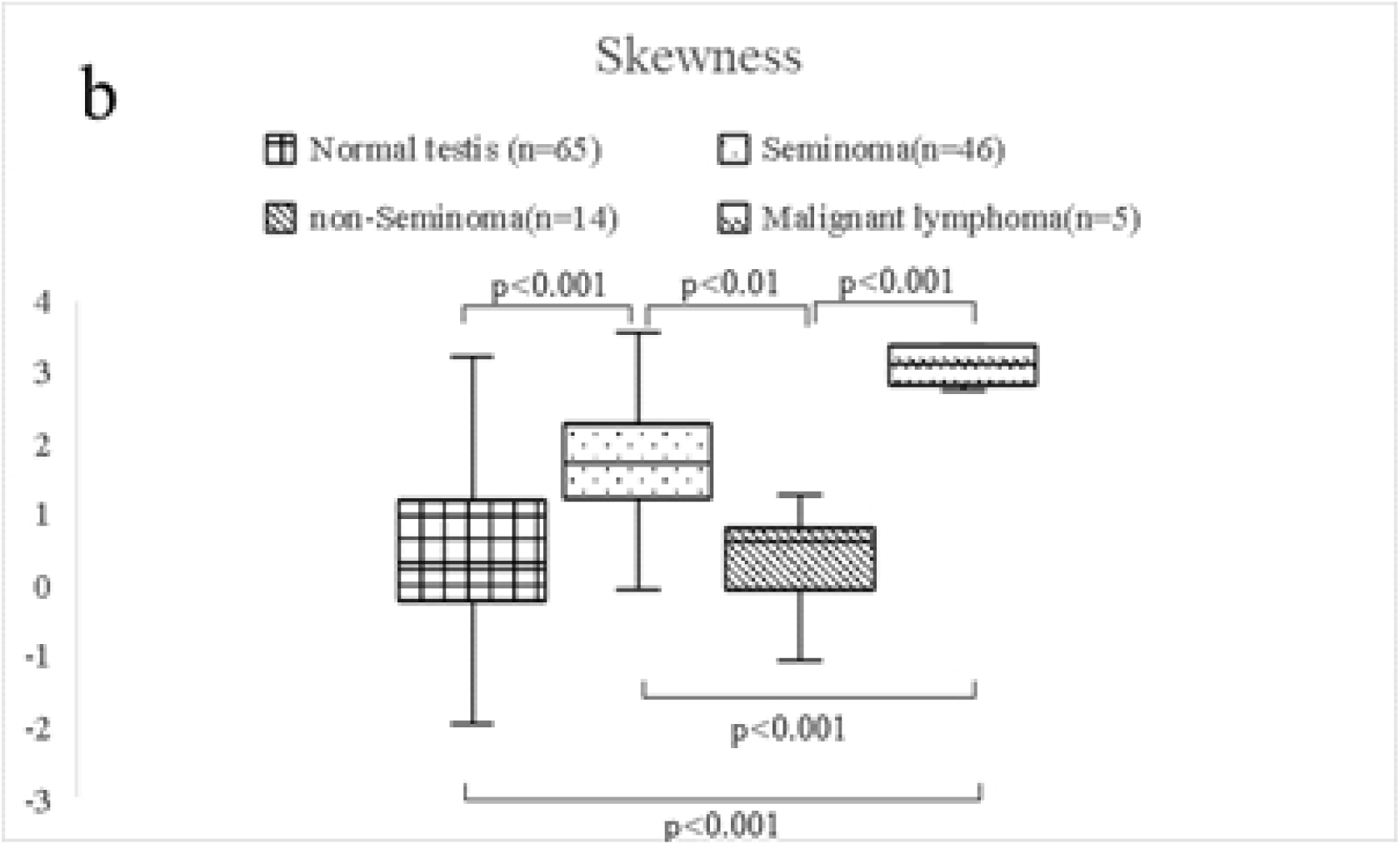

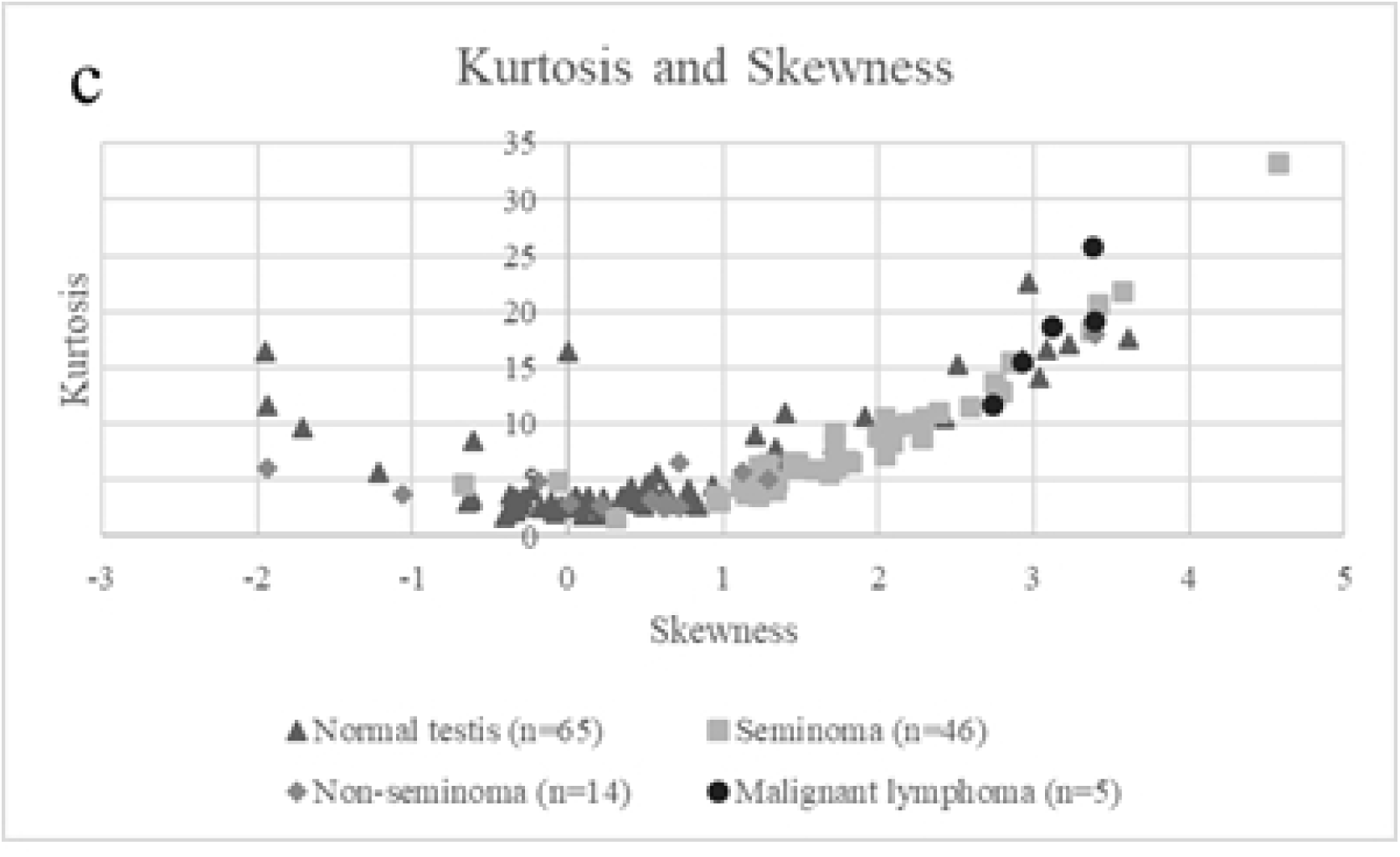

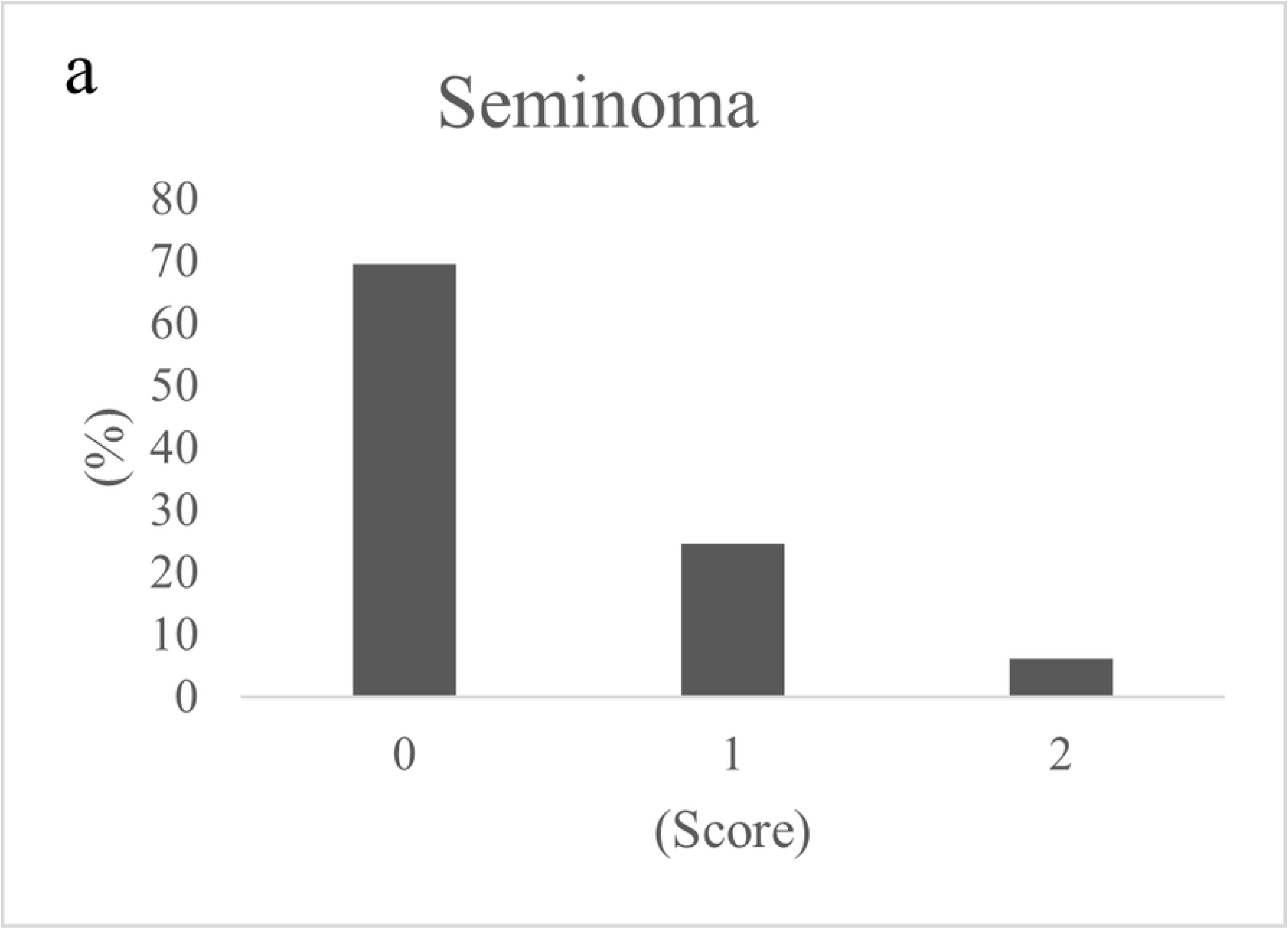

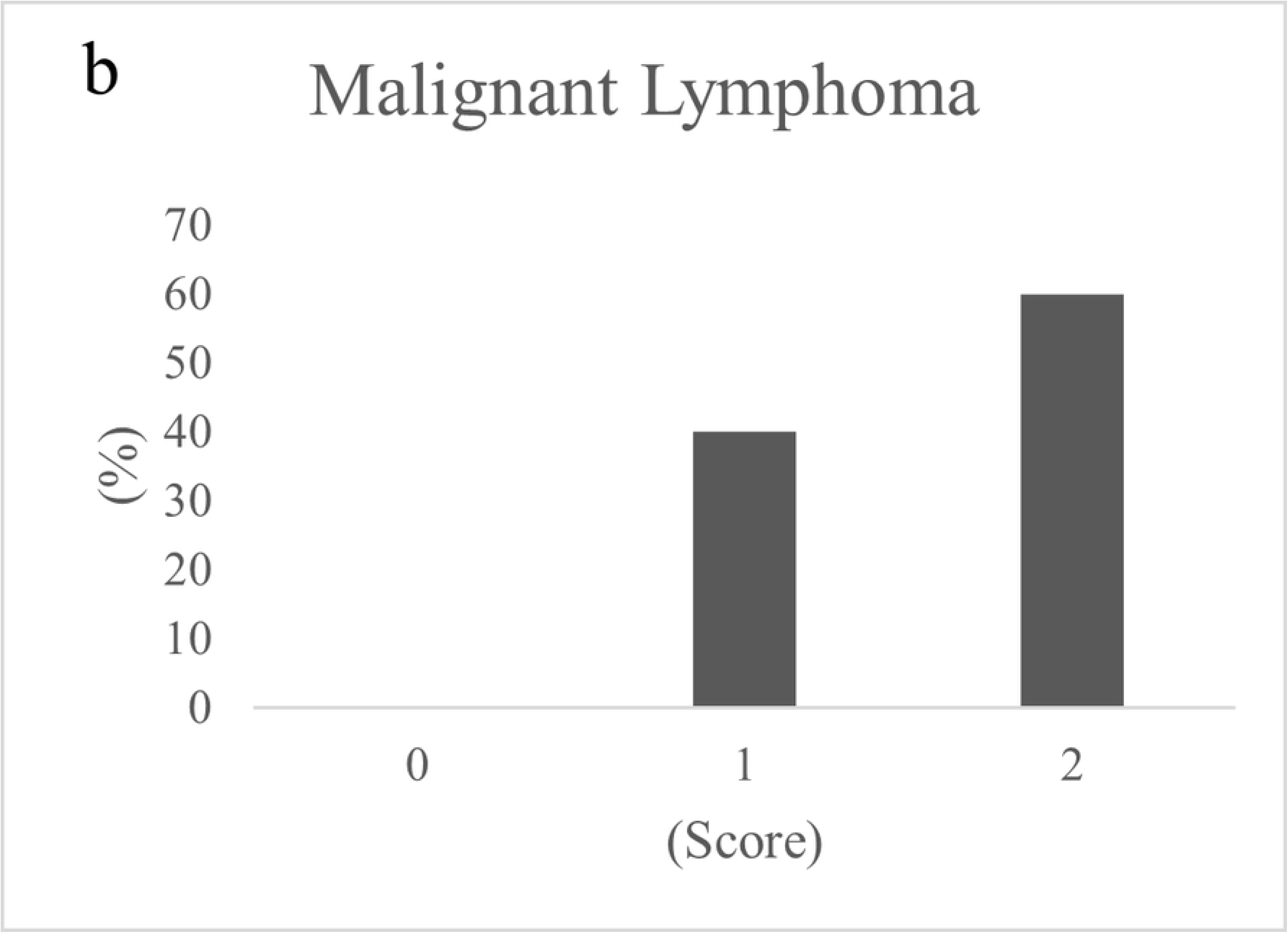

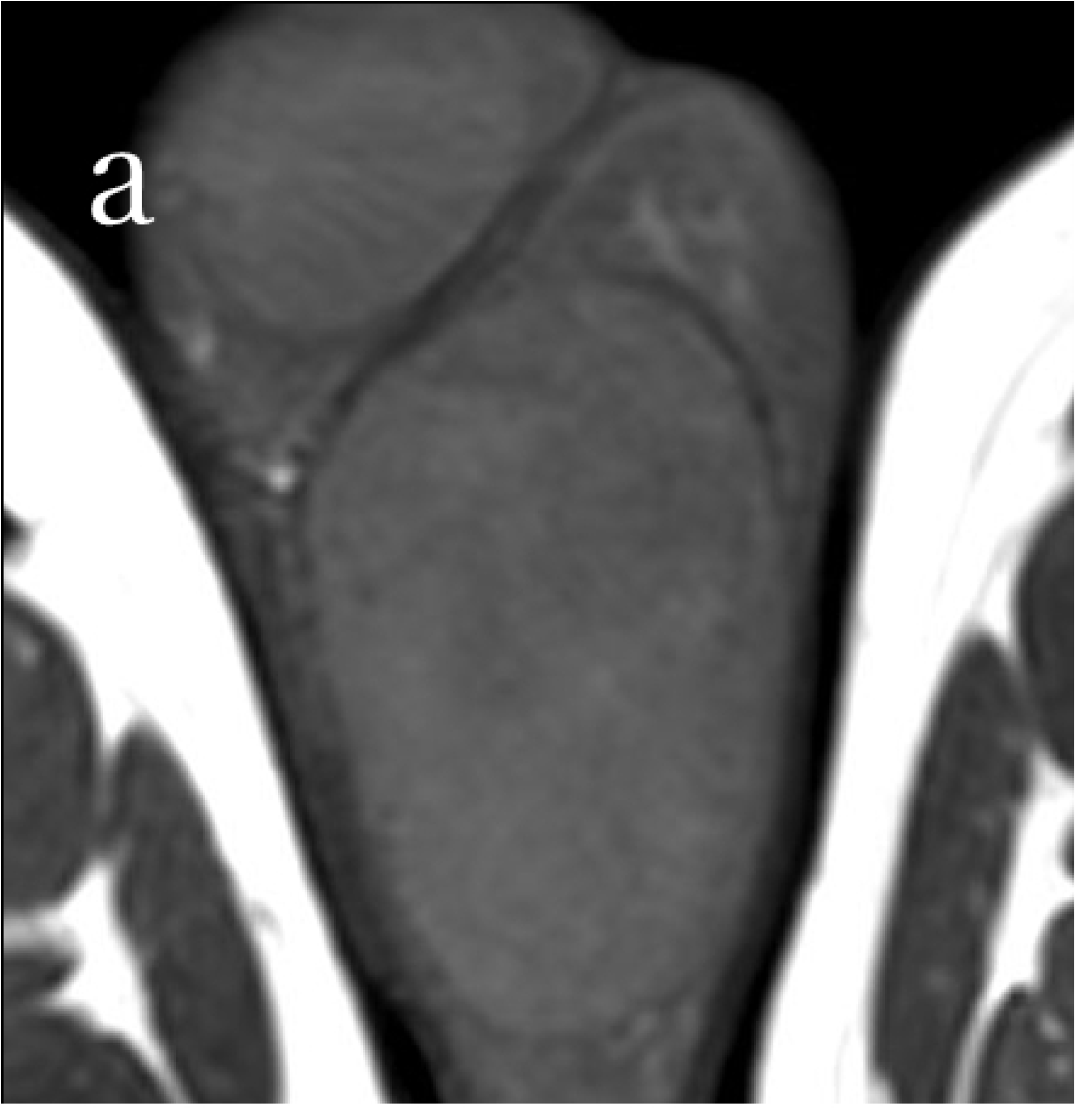

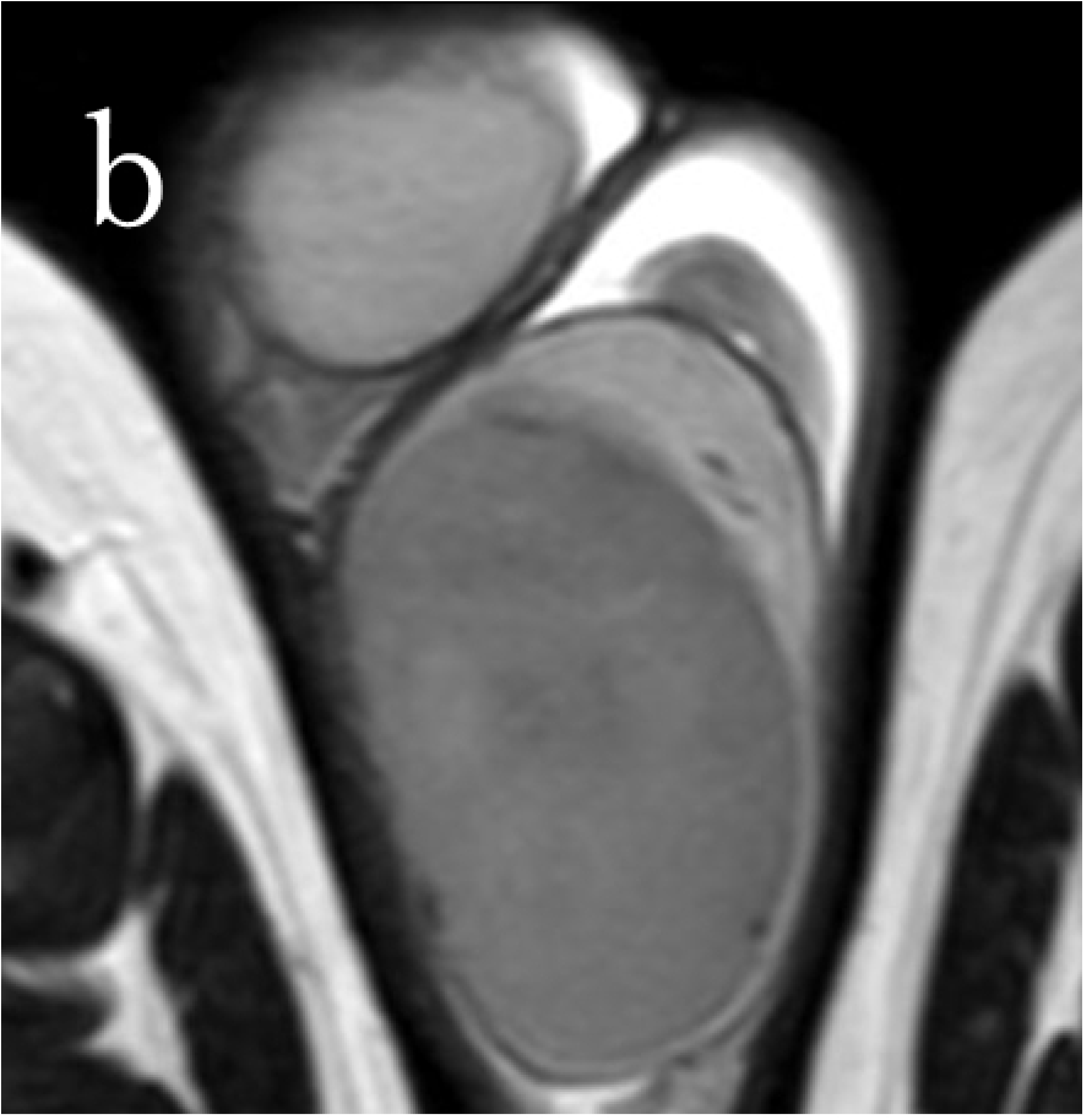

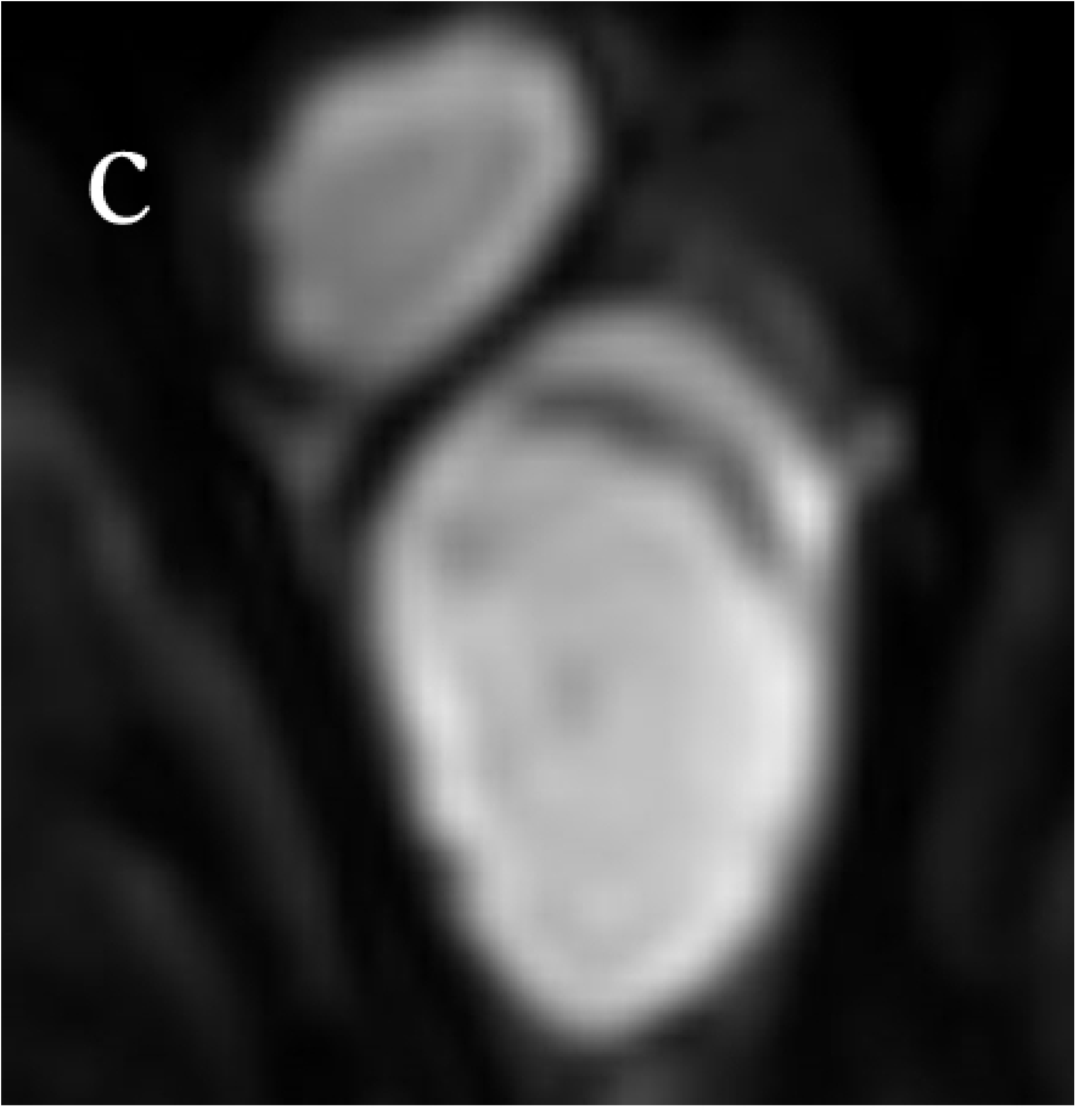

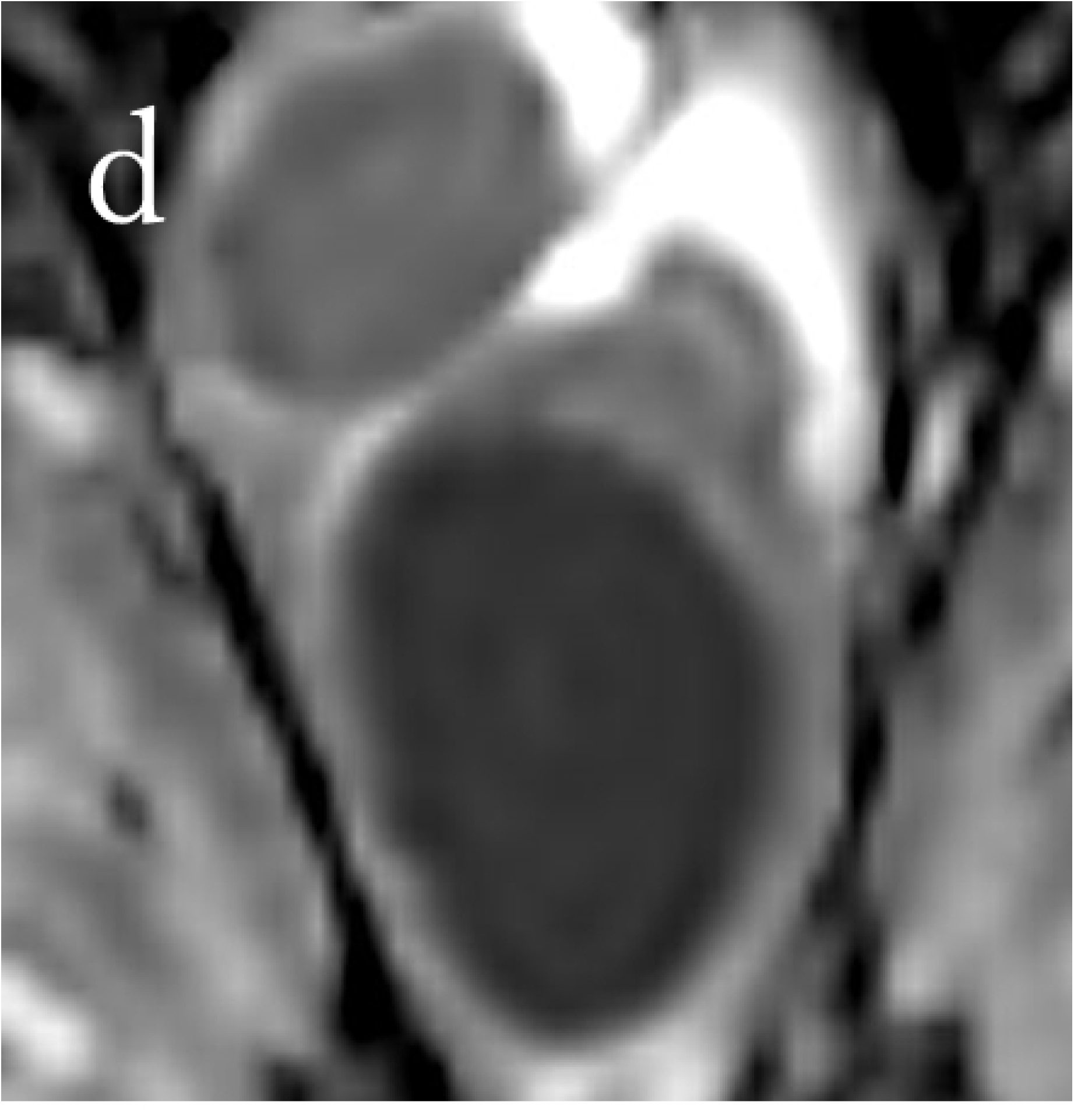

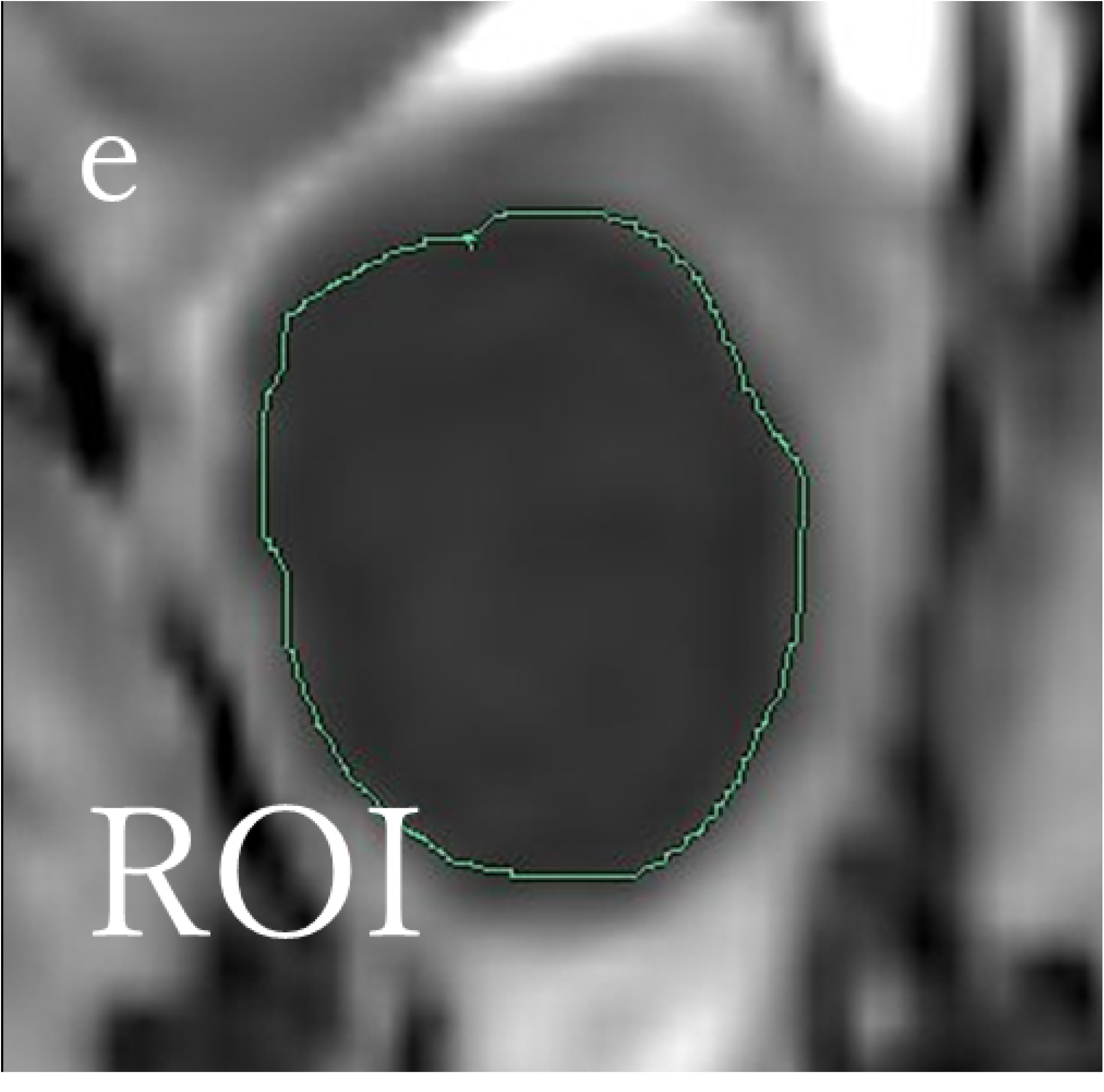

## Notes

### Competing Interest Statement

The authors have declared no competing interest.

### Funding Statement

The author(s) received no specific funding for this work.

### Author Declarations

the Bioethics Committee Dokkyo Medical University Saitama Medical Center

## References

1. Fonseca R, Habermann TM, Colgan JP, O’Neill BP, White WL, Witzig TE, et al. Testicular lymphoma is associated with a high incidence of extranodal recurrence. Cancer. 2000;88: 154–161. doi: 10.1002/(sici)1097-0142(20000101)88:1<154::aid-cncr21>3.0.co;2-t.

2. Tsili AC, Tsampoulas C, Giannakopoulos X, Stefanou D, Alamanos Y, Sofikitis N, et al. MRI in the histologic characterization of testicular neoplasms. AJR Am J Roentgenol. 2007;189: W331–W337. doi: 10.2214/AJR.07.2267.

3. Tanimoto A, Nakashima J, Kohno H, Shinmoto H, Kuribayashi S. Prostate cancer screening: the clinical value of diffusion-weighted imaging and dynamic MR imaging in combination with T2-weighted imaging. J Magn Reson Imaging. 2007;25: 146–152. doi: 10.1002/jmri.20793.

4. Lim HK, Kim JK, Kim KA, Cho KS. Prostate cancer: apparent diffusion coefficient map with T2-weighted images for detection-a multireader study. Radiology. 2009;250: 145–151. doi: 10.1148/radiol.2501080207.

5. Fütterer JJ, Heijmink SW, Scheenen TW, Veltman J, Huisman HJ, Vos P, et al. Prostate cancer localization with dynamic contrast-enhanced MR imaging and proton MR spectroscopic imaging. Radiology. 2006;241: 449–458. doi: 10.1148/radiol.2412051866.

6. Kitajima K, Kaji Y, Fukabori Y, Yoshida K, Suganuma N, Sugimura K. Prostate cancer detection with 3 T MRI: comparison of diffusion-weighted imaging and dynamic contrast-enhanced MRI in combination with T2-weighted imaging. J Magn Reson Imaging. 2010;31: 625–631. doi: 10.1002/jmri.22075.

7. Kim CK, Park BK, Lee HM. Prediction of locally recurrent prostate cancer after radiation therapy: incremental value of 3T diffusion-weighted MRI. J Magn Reson Imaging. 2009;29: 391–397. doi: 10.1002/jmri.21645.

8. Kunimatsu N, Kunimatsu A, Miura K, Mori I, Nawano S. Differentiation between solitary fibrous tumors and schwannomas of the head and neck: an apparent diffusion coefficient histogram analysis. Dentomaxillofac Radiol. 2019;48: 20180298. doi: 10.1259/dmfr.20180298.

9. Sun YS, Cui Y, Tang L, Qi LP, Wang N, Zhang XY, et al. Early evaluation of cancer response by a new functional biomarker: apparent diffusion coefficient. AJR Am J Roentgenol. 2011;197: W23–W29. doi: 10.2214/AJR.10.4912.

10. Lambregts DM, Beets GL, Maas M, Curvo-Semedo L, Kessels AGH, Thywissen T, et al. Tumour ADC measurements in rectal cancer: effect of ROI methods on ADC values and interobserver variability. Eur Radiol. 2011;21: 2567–2574. doi: 10.1007/s00330-011-2220-5.

11. Woo S, Cho JY, Kim SY, Kim SH. Histogram analysis of apparent diffusion coefficient map of diffusion-weighted MRI in endometrial cancer: a preliminary correlation study with histological grade. Acta Radiol. 2014;55: 1270–1277. doi: 10.1177/0284185113514967.

12. Gibbs P, Liney GP, Pickles MD, Zelhof B, Rodrigues G, Turnbull LW. Correlation of ADC and T2 measurements with cell density in prostate cancer at 3.0 Tesla. Invest Radiol. 2009;44: 572–576. doi: 10.1097/RLI.0b013e3181b4c10e.

13. Bakir VL, Bakir B, Sanli S, Yildiz SO, Iyibozkurt AC, Kartal MG, et al. Role of diffusion-weighted MRI in the differential diagnosis of endometrioid and non-endometrioid cancer of the uterus. Acta Radiol. 2017;58: 758–767. doi: 10.1177/0284185116669873.

14. Mimura R, Kato F, Tha KK, Kudo K, Konno Y, Oyama-Manabe N, et al. Comparison between borderline ovarian tumors and carcinomas using semi-automated histogram analysis of diffusion-weighted imaging: focusing on solid components. Jpn J Radiol. 2016;34: 229–237. doi: 10.1007/s11604-016-0518-6.

15. Kim YJ, Kim SH, Lee AW, Jin MS, Kang BJ, Song BJ. Histogram analysis of apparent diffusion coefficients after neoadjuvant chemotherapy in breast cancer. Jpn J Radiol. 2016;34: 657–666. doi: 10.1007/s11604-016-0570-2.

16. Schob S, Meyer HJ, Pazaitis N, Schramm D, Bremicker K, Exner M, et al. ADC histogram analysis of cervical cancer Aids detecting lymphatic metastases-a preliminary study. Mol Imaging Biol. 2017;19: 953–962. doi: 10.1007/s11307-017-1073-y.

17. Kurokawa R, Baba A, Kurokawa M, Capizzano A, Hassan O, Johnson T, et al. Pretreatment ADC histogram analysis as a prognostic imaging biomarker for patients with recurrent glioblastoma treated with bevacizumab: A systematic review and meta-analysis. AJNR Am J Neuroradiol. 2022;43: 202–206. doi: 10.3174/ajnr.A7406.

18. Umanodan T, Fukukura Y, Kumagae Y, Shindo T, Nakajo M, Takumi K, et al. ADC histogram analysis for adrenal tumor histogram analysis of apparent diffusion coefficient in differentiating adrenal adenoma from pheochromocytoma. J Magn Reson Imaging. 2017;45: 1195–1203. doi: 10.1002/jmri.25452.

19. Pope WB, Mirsadraei L, Lai A, Eskin A, Qiao J, Kim HJ, et al. Differential gene expression in glioblastoma defined by ADC histogram analysis: relationship to extracellular matrix molecules and survival. AJNR Am J Neuroradiol. 2012;33: 1059–1064. doi: 10.3174/ajnr.A2917.

20. Vitolo U, Ferreri AJ, Zucca E. Primary testicular lymphoma. Crit Rev Oncol Hematol. 2008;65: 183–189. doi: 10.1016/j.critrevonc.2007.08.005.

21. Sharbidre KG, Lockhart ME. Imaging of scrotal masses. Abdom Radiol (NY). 2020;45: 2087–2108. doi: 10.1007/s00261-019-02395-4.

22. Akyüz M, Topaktaş R, Ürkmez A, Koca O, Öztürk Mİ. Evaluation of germ-cell neoplasia in situ entity in testicular tumors. Turk J Urol. 2019;45: 418–422. doi: 10.5152/tud.2018.48855.

